# Multimodal Hypersensitivity Derived from Quantitative Sensory Testing Predicts Long-Term Pelvic Pain Outcome

**DOI:** 10.1101/2022.04.01.22272964

**Authors:** Matthew J. Kmiecik, Frank F. Tu, Daniel J. Clauw, Kevin M. Hellman

## Abstract

Multimodal hypersensitivity (MMH)—greater sensitivity across multiple sensory modalities (e.g., light, sound, temperature, pressure)—is hypothesized to be responsible for the development of chronic pain and pelvic pain. However, previous studies of MMH are restricted given their reliance on biased self-report questionnaires, limited use of multimodal quantitative sensory testing (QST), or limited follow-up. Therefore, we conducted multimodal QST on a cohort of 200 reproductive age women at elevated risk for developing or maintaining chronic pelvic pain conditions and pain-free controls. Pelvic pain self-report was examined over a four-year follow-up period. Multimodal QST was comprised of visual, auditory, bodily pressure, pelvic pressure, thermal, and bladder testing. A principal component analysis of QST measures resulted in three orthogonal factors that explained 43% of the variance: MMH, pressure stimulus-response, and bladder hypersensitivity. MMH and bladder hypersensitivity factors correlated with baseline self-reported menstrual pain, genitourinary symptoms, depression, anxiety, and health. Baseline self-report pain ratings were significant predictors of pelvic pain up to three years after assessment but decreased in their predictive ability of pelvic pain outcome over time. In contrast, MMH increased its predictive ability of pelvic pain outcome over time and was the only factor to predict outcome up to four years later. These results suggest that a “centralized” component of MMH is an important long-term risk factor for pelvic pain. Further research on the modifiability of MMH could provide options for future treatment avenues for chronic pain.

## 1 Introduction

Multimodal hypersensitivity (MMH) is a hallmark feature of chronic pain conditions (Curatolo et al., 2006; Fitzcharles et al., 2021; Geisser et al., 2008; Harper et al., 2016; Hollins et al., 2009; Sandri et al., 2021; Wilbarger & Cook, 2011) and may be a key reason why individuals develop chronic pain (Bar-Shalita et al., 2019; Greenspan et al., 2013) or a determinant of treatment response (Georgopoulos et al., 2019; Harte et al., 2016). MMH, also known as generalized sensory sensitivity, is increased sensitivity across multiple sensory modalities (e.g., light, sound, temperature, pressure) common in “centralized” pain conditions (Arendt-Nielsen, 2017; Fitzcharles et al., 2021; Schrepf et al., 2018). Individuals with chronic pelvic pain conditions, like irritable bowel syndrome and bladder pain syndrome, similarly exhibit MMH despite not having any outward pelvic pathology (e.g., infection, endometriosis, etc.; Kaya et al., 2013; Schrepf et al., 2018). Previous studies have attempted to quantify MMH (Aykan et al., 2020; Brown et al., 2001; Greenspan et al., 2011; Lionetti et al., 2018), its stability over time (Schrepf et al., 2018), and its relationship to chronic pain severity (Greenspan et al., 2020; López-Solá et al., 2014) and outcome (Harte et al., 2019; Morris et al., 2021; Müller et al., 2021; Slade et al., 2014); however, our understanding of MMH is limited given these investigations often relied on subjective self-report questionnaires, inadequately assessed MMH by using uni-dimensional quantitative sensory testing (QST), or lacked long-term follow-up. Despite the ubiquitous use of QST to understand pain conditions (Treede, 2019), there is limited understanding of how different QST methods relate to each other and predict changes in pelvic pain symptomatology (Grundström et al., 2019; Morris et al., 2021). A synergistic approach encompassing multiple modalities of QST with longitudinal symptom assessment could improve our understanding of MMH, further quantify an individual’s susceptibility to developing chronic pain, and accelerate the development of preventative measures and treatments.

Therefore, we performed secondary analyses on a four-year longitudinal cohort of young reproductive age women that had multimodal QST assessed at baseline (CRAMPP: Chronic Pain Risk Associated with Menstrual Pelvic Pain; NCT02214550). Because menstrual pain is among the leading risk factors for chronic pelvic pain (Li et al., 2020; Zondervan et al., 2001), we focused recruitment of women with menstrual pain. We also included pain-free controls and a subset of women with chronic pelvic pain to provide a full range of sensory profiles for analysis. CRAMPP’s multimodal QST battery was comprised of provocation with pressure, cold, and audio/visual stimuli, temporal summation (a measure of spinal wind-up), conditioned pain modulation (a metric of descending inhibition), and bladder distension (a measure of visceral sensitivity). Analysis of CRAMPP’s baseline data revealed that individuals with dysmenorrhea and bladder pain hypersensitivity have impaired conditioned pain modulation, and increased sensitivity to pressure and thermal (i.e., cold) (Hellman et al., 2020), and visual stimuli (Kmiecik et al., 2021), even without any formal chronic pain diagnoses. However, we have not explored whether these QST changes were unified (i.e., MMH) and whether altered QST predicted long-term pelvic pain outcome. Therefore, we analyzed the measures obtained from sensory testing in two ways: 1) In an effort to understand which sensory testing procedures predicted future pelvic pain, we combined our testing into three constructs: traditional QST, non-invasive bladder distension (i.e., provoked visceral sensitivity), and supraspinal audio/visual sensitivity; and 2) we reduced the dimensionality of our QST data using principal component analysis (PCA) to characterize the sources of underlying multimodal QST variability and identify a unified MMH construct. We hypothesized that MMH would predict long-term pelvic pain outcome and outperform uni-dimensional constructs of sensory testing (i.e., QST, bladder testing, audio/visual sensitivity).

## 2 Methods

A brief Methods section is reported below. Readers are referred to the Supplemental Materials for more detailed descriptions across the Participants, Procedure, and Statistical Analyses sections.

### 2.1 Participants

A total of 354 participants were enrolled in CRAMPP. A full enrollment diagram is reported elsewhere (Hellman et al., 2020). All participants provided informed consent and followed protocols approved by NorthShore University HealthSystem’s Institutional Review Board. Participants were monetarily compensated for their time.

Enrolled participants included women with low menstrual pain (≤3 on a 0-10 Numerical Rating Scale [NRS]; 0 = no pain; 10 = worst pain imaginable) or moderate to severe menstrual pain (≥5 on a 0-10 NRS) but no other chronic pain, those diagnosed with bladder pain syndrome (BPS), and those diagnosed with a non-pelvic chronic pain condition (general pain ≥5 on a 0-10 NRS for more than three consecutive months). BPS participants were required to meet American Urological Association diagnostic criteria, and report bladder pain ≥3 on a 0-10 NRS for more than 3 consecutive months (Hanno et al., 2011).

From these 354 women, a total of 200 participants had complete QST data from their baseline assessment visit. Participants completed annual questionnaires following their baseline visit for up to five years. Given the ongoing collection of year five annual questionnaires (*n*=63), we included here completed annual questionnaires up to year four to increase our statistical power (*n*=87). Demographic variables of interest are presented in Table 1.

**Table 1:** Participant Demographics. *Note*. Percentages were calculated from total sample size (*n*=200). Some participants had multiple diagnoses.

| Measure | $n$ (%) | Mean ($SD$) | Measure | $n$ (%) |
| --- | --- | --- | --- | --- |
| **Race** |  |  | **Prior Health Diagnoses** |  |
| White | 121 (60.5%) |  | Ovarian Cysts | 27 (14%) |
| Black or African American | 32 (16%) |  | Lower Back Pain | 21 (11%) |
| Asian | 30 (15%) |  | Chronic Pelvic Pain | 18 (9%) |
| Multiple | 15 (7.5%) |  | Migraine Headaches | 18 (9%) |
| Native American | 1 (.5%) |  | Irritable Bowel Syndrome | 16 (8%) |
| No Response | 1 (.5%) |  | Bladder Pain Syndrome | 13 (7%) |
| **Ethnicity** |  |  | Endometriosis | 13 (7%) |
| Not Hispanic or Latino | 176 (88%) |  | Chronic Constipation | 6 (3%) |
| Hispanic or Latino | 24 (12%) |  | Fibroids | 5 (3%) |
| **Groups** |  |  | Inflammatory Bowel Disease | 5 (3%) |
| Dysmenorrhea | 132 (66%) |  | Kidney Stones | 4 (2%) |
| Pain-Free Controls | 30 (15%) |  | Fibromyalgia | 3 (2%) |
| Chronic Pain | 22 (11%) |  | Pelvic Inflammatory Disease | 1 (1%) |
| Bladder Pain Syndrome | 16 (8%) |  | Chronic Diarrhea | 1 (1%) |
| **Age (years)** | 200 | 24.9 (6.4) | Arthritis | 1 (1%) |
| **Height (inches)** | 196 | 64.6 (2.8) | **Parity** |  |
| **Weight (lbs.)** | 195 | 143 (30.1) | Ever Pregnant | 29 (15%) |
| **Body Mass Index (BMI)** | 195 | 24 (4.63) | 1 or more deliveries | 14 (7%) |

### 2.2 Procedure

Eligible participants that were enrolled in the study first participated in a screen visit and a second, baseline assessment visit. In the baseline visit, which was performed during the participants’ midluteal (pain-free) phase of their menstrual cycle, participants completed a panel of questionnaires that assessed their health history. Participants next underwent a quantitative sensory testing (QST) panel that included mechanosensation, cold pressor, visceral provocation, conditioned pain modulation (CPM), and temporal summation (TS). All QST measures and self-report questionnaires are detailed in the Supplementary Materials and are briefly explained here. We measured bladder sensitivity using our validated non-invasive bladder filling test that measures urgency, pain, volume, and pain descriptors across a range of thresholds (Hellman et al., 2018; Tu et al., 2013; Tu et al., 2017). Body and pelvic pain sensitivity was assessed as pressure pain thresholds (PPTs) across four body and four transvaginal sites using a digital algometer (Wagner Instruments, Greenwich, CT) and finger mounted force-sensing resistor (Trossen Robotics, Downers Grove, IL), respectively (Hellman et al., 2015; Hellman et al., 2020). CPM was assessed by performing repeat PPT testing of the left knee before and after ice water immersion of the contralateral hand (Nir & Yarnitsky, 2015). To measure TS, we delivered 10 pressure pulses at the identified pain threshold to the right knee using a digital algometer and captured participant pain ratings after each pulse (Cathcart et al., 2009). Although not typically included in QST, we also assessed participants’ unpleasantness ratings during aversive visual and auditory stimulation. In the visual task, participants rated their experienced unpleasantness after viewing a periodic pattern-reversal blue/yellow checkerboard stimulus across five blocks of varying brightness intensities (Harte et al., 2016; Kmiecik et al., 2021). For the auditory task, participants similarly rated their experienced unpleasantness after listening to an aversive tone across five blocks of varying loudness intensities (Hollins et al., 2009).

#### 2.2.1 Pelvic Pain Outcome

Participants completed annual questionnaires virtually by email in REDCap for up to four years. As part of this questionnaire, participants rated their average feeling of 1) menstrual and non-menstrual pelvic pain, 2) pain with urination, and 3) pain with bowel movements during the past week using a 0-100 VAS (0=no pain; 100=worst pain imaginable). VAS scales are more sensitive to changes than descriptive word-based scales (Sriwatanakul et al., 1983) and have linear properties amenable to averaging (Myles et al., 1999). Therefore, we averaged these three questions to create a composite pelvic pain outcome variable. Similar composite pain recall variables formed by averaging have demonstrated high validity and reliability comparable to daily diary ratings of pain (Jensen et al., 2012).

#### 2.2.2 QST, Bladder Test, and Audio/Visual Predictor Constructs

To evaluate the ability of different sensory tests to predict pelvic pain outcome, we combined the 40 measures from the multimodal QST panel into three constructs using summed Z-scores: traditional QST measures, bladder test measures, and audio/visual stimulation. The QST construct comprised 25 measures including PPTs (i.e., thresholds, after pain, and descriptors), TS, CPM, and cold pain. The bladder test construct comprised 11 measures, including pain, urgency and descriptors. The audio/visual sensitivity construct comprised 4 measures, including mean unpleasantness and slope of the stimulus-response function. Prior to calculating Z-scores, all measures maintained the same directionality such that greater values denoted increased pain/impairment/sensitivity. Final constructs were mean centered for regression analyses.

### 2.3 Statistical Analyses

To assess how well baseline QST, bladder test, and audio/visual measures independently predicted future pelvic pain outcome, we performed four multiple regressions using self-report data acquired from annual follow-up questionnaires.

Pelvic pain outcome (defined in Section 2.2.1) served as the dependent variable. The independent variables included the summed Z-scores (QST, bladder test, audio/visual sensitivity) defined above (see Section 2.2.2). We also included baseline pelvic pain outcome as a covariate. All independent variables were measures collected at the participants’ baseline visit.

Additionally, we performed a principal components analysis (PCA) to better understand the factors underlying QST variability (Abdi & Williams, 2010). Our data matrix columns comprised the 40 measures derived from the multimodal QST panel (see Section 1.2.1). All measures maintained the same directionality such that greater values denoted increased pain/impairment/sensitivity/etc. Each column was then Z-scored prior to decomposing the matrix via singular value decomposition (SVD; Abdi, 2007).

Inferential statistics were performed using data resampling techniques (Abdi & Williams, 2010; Beaton et al., 2014). Permutation testing quantified probability values for observed principal components (PCs) and aided in their interpretation. Bootstrapping quantified each measure’s loading stability/contribution importance using bootstrap ratios (BSRs). A BSR is the ratio between a measure’s fixed-effect factor score (i.e., loading) and the standard deviation of its bootstrapped distribution and is interpreted like Student’s *t* value. Therefore, significantly contributing measures have |*BSRs*| > 1.96 (*p <* .05).

To compare QST-based PCs with measures not included in their initial formulation, we calculated bootstrapped correlations between the row-wise (i.e., participant) factor scores and validated self-report questionnaires (see Section 1.2.2). To assess how well QST-based PCs predicted future pelvic pain outcome, we repeated the multiple regression procedure as described above except that pertinent PCs served as the independent variables instead of the three Z-scored measures.

A sensitivity analysis was performed to examine whether adjusting for prevalence rates of dysmenorrhea, bladder pain syndrome, and other types of chronic pain in the general population altered regression results. Keeping our sample size constant (*n*=200), participants were sampled with replacement according to the following prevalence rates (Berry et al., 2011; Dahlhamer et al., 2018; Schoep et al., 2019): 50% pain-free healthy controls (*n*=100), 40% moderate-severe dysmenorrhea (*n*=80), 5% bladder pain syndrome (*n*=10), 5% other chronic pain (*n*=10). Regression analyses were recomputed using these bootstrapped data samples. This procedure was repeated for 2,000 iterations. Mean regression estimates and effect sizes were then compared to original un-adjusted values.

All code and data for this investigation is available on GitHub (https://github.com/mkmiecik14/mmh) and OSF (insert link here upon publication).

## 3 Results

### 3.1 QST and Visceral Sensitivity Modestly Predict Pelvic Pain Outcome

Distributions of predictor variables and pelvic pain outcome are visualized in Figure 1A and B, respectively (see Supplemental Table 1 and 2 for complete QST data). Cross-sectional relationships between sensory tests and baseline pelvic pain demonstrated that sensitivity across the QST, bladder test, or audio/visual constructs were associated with worse baseline pelvic pain (see Figure 1E).

**Figure 1:**
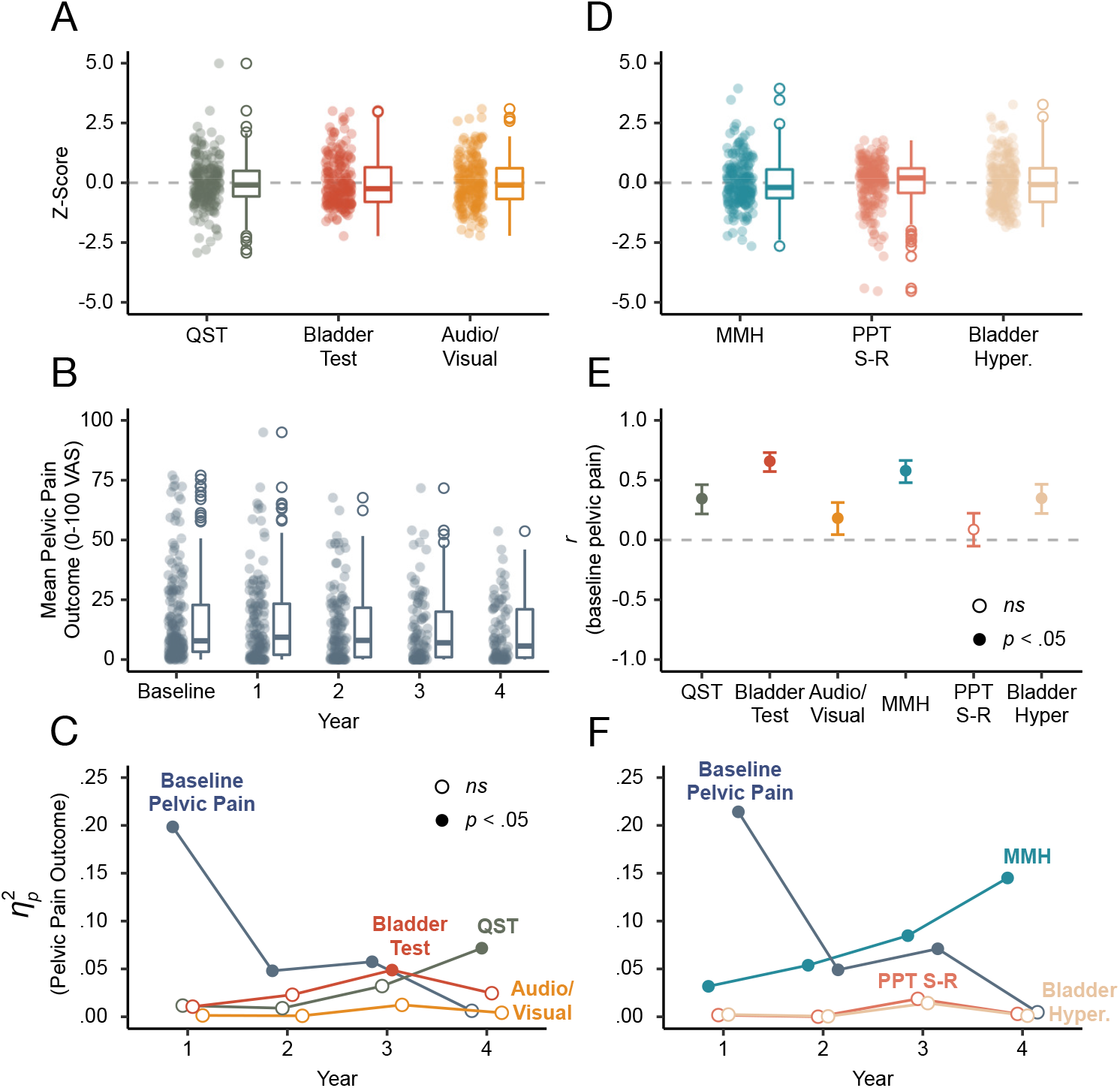
MMH is the best predictor of future pelvic pain. A) Box plots of Z-scores for each sensory testing construct. B) Box plots of pelvic pain outcome across the baseline visit and annual questionnaires. C) Explained variance 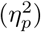 of pelvic pain outcome for each sensory testing construct and baseline pelvic pain. Significant predictors (filled circles) include baseline pelvic pain, bladder test, and QST. D) Box plots of PCs used for regression modeling. E) Correlations between baseline pelvic pain and predictors (i.e., sensory testing constructs and PCs). Error bars are 95% confidence intervals and all correlations were significant (filled circles) except PPT S-R. F) Explained variance 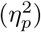 of pelvic pain outcome for each PC accounting for baseline pelvic pain. Significant predictors (filled circles) include MMH and baseline pelvic pain.

Multiple regressions were used to assess how well QST, bladder test, and audio/visual constructs predicted pelvic pain outcome on annual questionnaires administered up to four years following the baseline visit (see Supplementary Table 3). We accounted for baseline pain by including it as a covariate (see Supplementary Table 6 for descriptive statistics). Baseline pelvic pain was the strongest predictor of year 1 pelvic pain, but steadily declined over time (see Figure 1C). The bladder test and QST predicted pelvic pain at years 3 and 4, respectively. Audio/visual testing did not predict outcome at any year. After adjusting for population-based prevalence rates of patient groups, baseline pelvic pain was a stronger predictor of year four pelvic pain outcome (see Supplementary Figure 3). Also, QST explained little to no variance in pelvic pain outcome at any year. Thus, overall baseline pelvic pain was a better predictor of future pelvic pain than uni-dimensional QST, bladder test, and audio/visual constructs.

### 3.2 PCA Identifies Three Components Explaining QST Variability

We determined the number of principal components (PCs) underlying QST variability across the cohort by examining the scree plot (see Supplementary Figure 1), permutation testing results, and loadings using geometrically plotted factor scores of QST measures (Abdi and Williams, 2010; see Supplementary Materials Table 4). Accordingly, we identified three components as interpretable. The first PC (PC1) explained 20.6% of the variance (*p* = .0005), the second (PC2) 12.4% (*p* = .0005), and the third (PC3) 9.5% (*p* = .0005).

Factor score plots for the first three PCs are shown in Figure 2 (see Supplementary Figure 2 for bootstrapped significance of factor loadings). Given that all measures loaded positively on PC1, we interpret PC1 to represent MMH (i.e., increased sensitivity on one measure was associated with an increased sensitivity on another). Forehead, hip, knee, shoulder, and vaginal PPTs positively loaded on PC2, while their respective after-pain ratings were opposed on PC2. This factor is representative of a stimulus-response function of pressure and after-pain resulting from PPT testing, hereafter referred to as PPT S-R (PPT stimulus-response). In other words, participants with lower PPTs (i.e., less force, greater sensitivity) reported less after-pain ratings. PC3 depicted an opposing relationship between bladder task measures and PPTs (thresholds and after-pain ratings). Given the orthogonality of PCs, PC3 captured bladder pain hypersensitivity that was distinct from MMH (PC1).

**Figure 2:**
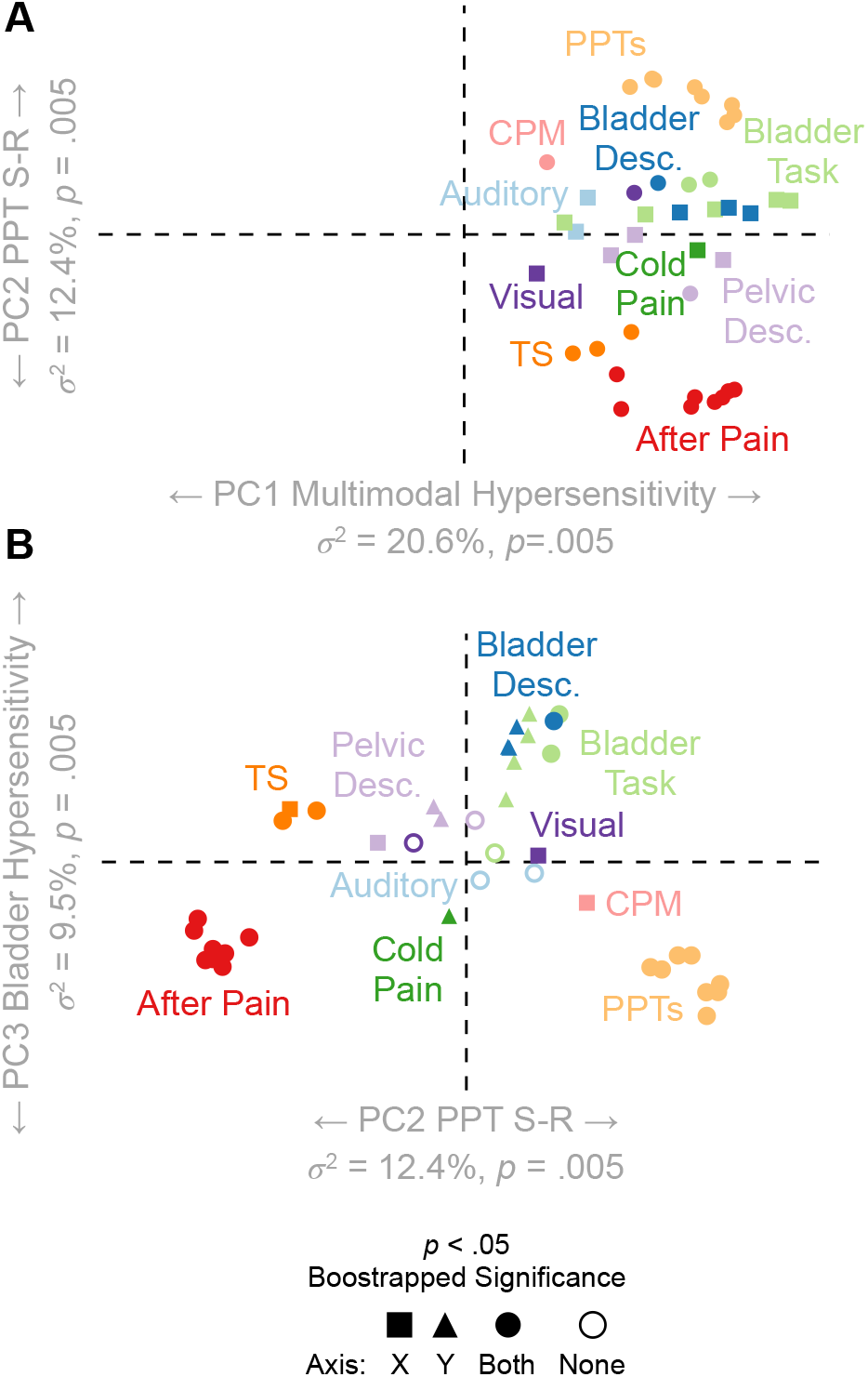
Factor Score Plots of QST Variables. Each measure’s position denotes its relative loading/correlation with the orthogonal principal components (PCs) plotted across the x-y coordinate plane. Measures in close proximity within an axis plane depict positive relationships (concomitant sensitivity across proximal measures). Measures that are distant appear on opposite sides of the origin and depict negative relationships within an axis plane. A boot-strapping procedure quantified loading significance for each measure on each PC depicted using shapes. A) Within the x-axis plane (PC1), all QST variables load positively onto PC1 and are proximal, indicating positive relationships across all QST measures. B) Within the x-axis plane (PC2), PPTs and after-pain measures are distant from each other and are located on opposite sides of the origin. This indicates a negative relationship between PPTs and after-pain measures (this can also be seen across the y-axis in A). Within the y-axis plane (PC3), measures from the bladder task positively load onto PC3 and are distant/oppose the PPTs and after-pain measures. PC=Principal Component; PPTs=Pressure Pain Thresholds; CPM=Conditioned Pain Modulation; TS=Temporal Summation.

### 3.3 PCs Correlate with Baseline Self-Report Measures

Figure 3 depicts the bootstrapped correlations between row-wise factor scores (i.e., participants) of PCs and validated questionnaires of self-reported menstrual pain, genitourinary symptoms, depression, anxiety, and health (see Supplementary Table 5 for descriptive statistics of self-report measures). PC1 (MMH) and PC3 (bladder hypersensitivity) correlated strongly with every measure included, while PC2 (PPT S-R) only weakly correlated with the Interstitial Cystitis Symptom Inventory (ICSI) and Genitourinary Pain Index (GUPI), two standardized clinical questionnaires for bladder pain. These widespread correlations observed across PCs 1 and 3, but not PC2, demonstrate that these two dimensions (MMH and bladder pain hypersensitivity) explain variability in participants’ current pain- and health-related quality of life. Also, given the orthogonality of PCs, these results suggest the contribution of two mechanisms to explain patients’ current pelvic pain health-related quality of life : 1) MMH and 2) bladder hypersensitivity.

**Figure 3:**
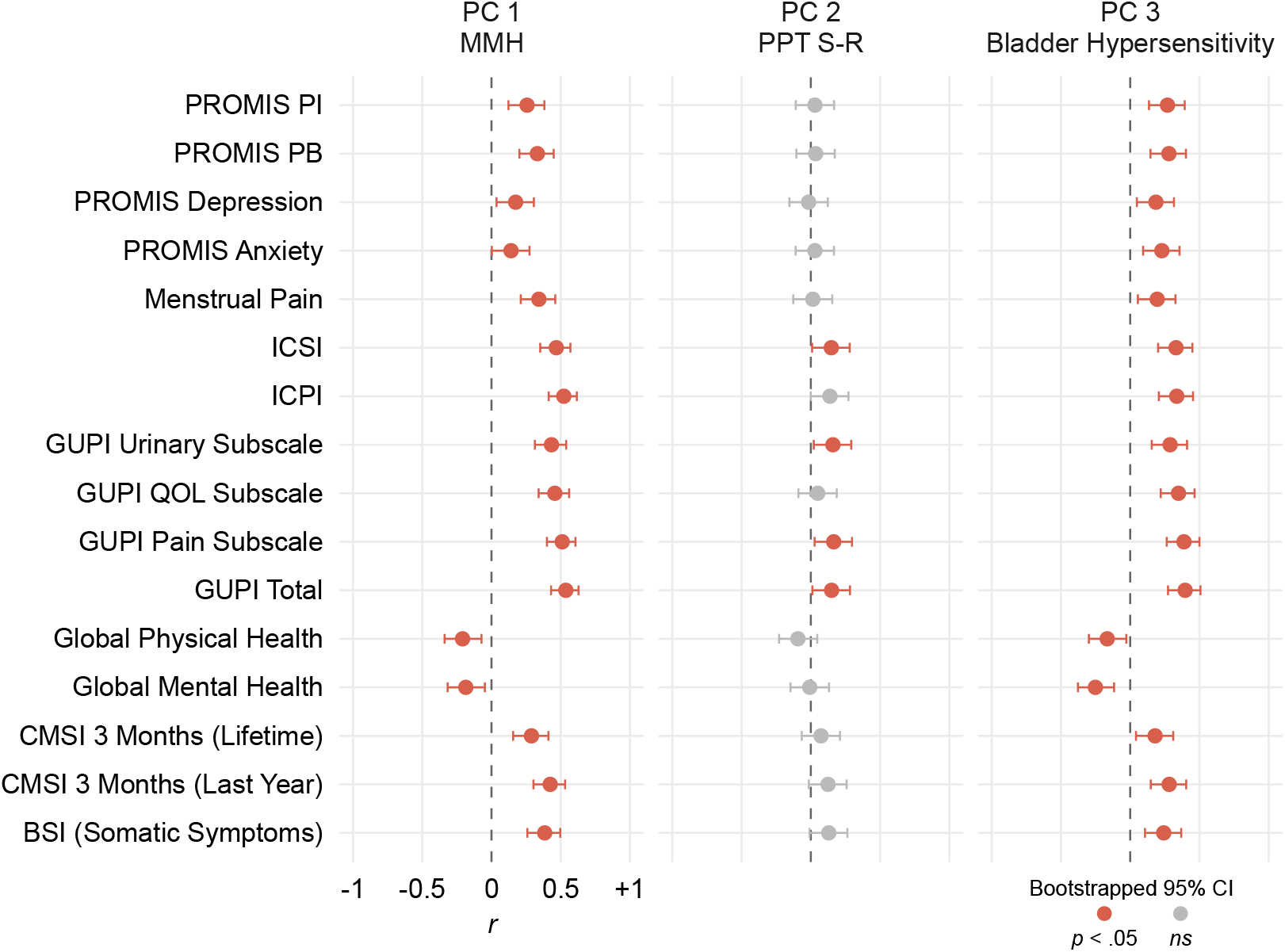
Correlations Between Principal Components (PCs) and Self-Report Questionnaires. Each point depicts a Pearson’s pairwise correlation between the participants’ factor scores across the three PCs of interest and their responses to a self-report questionnaire. Error bars denote boot-strapped 95% confidence intervals; therefore, intervals crossing zero indicate non-significant correlations and are colored grey. Except in the case of global mental/physical health, greater scores on self-report questionnaires denote worse symptoms/outcome. All MMH and bladder hypersensitivity correlations were significant and in the expected direction: greater hypersensitivity was associated with worse affective symptoms, somatic symptoms, genitourinary pain, menstrual pain, and overall pain and health.

### 3.4 Baseline MMH Predicts Longitudinal Pelvic Pain Outcome Four Years Later

Multiple regressions were used to assess how well the three obtained PCs (i.e., MMH, PPT S-R, and bladder hypersensitivity) predicted pelvic pain outcome on annual questionnaires administered up to four years following the baseline visit (see Supplementary Table 7). Distributions of the PCs are presented in Figure 1D. Similar to the self-report measures, baseline pelvic pain correlated with both MMH and bladder hypersensitivity, but not with PPT S-R (see Figure 1E). PCs were orthogonal to each other (*r* =0).

Baseline pelvic pain was the strongest predictor of year 1 pelvic pain, but steadily declined over time. In contrast, MMH (PC1) increased in its predictability of pelvic pain outcome over time and predicted worse pelvic pain outcome continuously up to four years later (see Figure 1F). A 1*SD* increase in MMH at baseline predicted a .44*SD* increase, or nearly 6 VAS points, in pelvic pain ratings four years later. PPT S-R (PC2) and bladder pain hypersensitivity (PC3) did not predict outcome at any year. Adjusting for population-based prevalence rates of included patient groups replicated the observed sample-wise regression results (see Supplementary Figure 3). Thus, the positive association between MMH and pelvic pain was robust and differentiated participant four-year trajectories (see Supplemental Figure 4).

## 4 Discussion

Understanding mechanisms underlying pain sensitivity, and its prospective risk for future pain, remains a hurdle towards progress in treating and preventing chronic pain conditions. QST is the most widely used method for systematically measuring pain sensitivity (Cruz-Almeida & Fillingim, 2014; Curatolo, 2011; Rolke et al., 2006); however, traditional assays that measure single nociceptive modalities (e.g., thermal, pressure) have demonstrated inconsistent predictive power for pain outcome (Bordeleau et al., 2021; Forstenpointner et al., 2021; Schmelz, 2020; Treede, 2019). Likewise, the ability of dynamic QST paradigms assessing pain modulation, such as CPM and TS, to predict pain outcome have also been mixed (Fernandes et al., 2019; Morris et al., 2021; Nir & Yarnitsky, 2015; O’Brien et al., 2018). Our results demonstrate that the utility of QST can be improved by measuring sensory hypersensitivity more broadly (i.e., including several disparate sensory modalities to evaluate MMH). This approach is aligned with the repeated observation that many functional pain syndromes additionally report increased sensitivity to environmental stimuli (e.g., lights, sounds, odors). Thus, dysfunction in global sensory processing may be more crucial to chronic pain risk than any nociceptive mechanism underlying hyperalgesia.

In a cohort of women with a range of chronic pelvic pain risk, we found that including both nociceptive and non-nociceptive (e.g., visual and auditory) assays identified that MMH was a robust common denominator underlying QST variability. Importantly, baseline MMH predicted pelvic pain annually for four years even when accounting for baseline pelvic pain. MMH, rather than solitary dysfunction in specific nociceptive modalities or pain modulation, appears to underlie pain vulnerability and likely plays a substantial role in the development of chronic pain conditions.

### 4.1 MMH: Nociceptive and Non-Nociceptive QST

QST studies often infer centralized or generalized mechanisms of pain sensitivity to explain observed hypersensitivity in chronic pain conditions (e.g., FitzGerald et al., 2005; Geisser et al., 2008; Greenspan et al., 2020; Grundström et al., 2019; Harte et al., 2016; Harte et al., 2019; Hellman et al., 2020; Hollins et al., 2009; Kmiecik et al., 2021; Martenson et al., 2016). Examining relationships between and within QST modalities has provided support that QST assays measure distinct mechanisms of pain sensitivity (Greenspan et al., 2011; Hastie et al., 2005; Janal et al., 1994; Lautenbacher & Rollman, 1993). However, given that most studies relied on singular noxious modalities of QST, or combinatory approaches of select modalities (e.g., heat and pressure), they provide only a limited ability to evaluate generalized mechanisms of hypersensitivity in centralized pain conditions (Bhalang et al., 2005).

The current investigation improves upon these previous attempts by administering a broader panel of nociceptive sensory tests (PPTs, CPM, TS, cold pressor, bladder provocation) and non-nociceptive supraspinal tests (visual and auditory sensitivity). Like previous QST studies (Greenspan et al., 2011; Hastie et al., 2005), a PCA of these 40 QST measures resulted in modality-specific components: PPT S-R (PC2; similar to Santana et al., 2020) and bladder hypersensitivity (PC3; similar to Tu et al., 2017). In contrast, the largest source of underlying QST variability was MMH, a component that was not modality specific. Although previous studies have conjectured that centralized hypersensitivity (e.g., generalized sensory sensitivity, somatization, somatic symptoms disorders, sensory modulation disorder, etc. Bar-Shalita et al., 2019; Birket-Smith, 2001; Curatolo et al., 2006; Schrepf et al., 2018) underlies chronic pain conditions, our results provide experimental evidence of MMH being a broad construct that correlates with affective symptoms, somatic symptoms, genitourinary pain, menstrual pain, and overall pain and health. Given the consistent and strong correlation between MMH and these other factors crucially affecting chronic pain, we hypothesize neural mechanisms underlying MMH are critical circuits in functional pain syndromes.

### 4.2 MMH Predicts Pelvic Pain Outcome

Studies that have used various QST measures to predict long-term outcome have not generated uniform conclusions (e.g., Fernandes et al., 2019; Georgopoulos et al., 2019; Greenspan et al., 2013; Harte et al., 2019; Morris et al., 2021; Müller et al., 2021; Slade et al., 2014; Yarnitsky et al., 2008; Yarnitsky et al., 2012). In contrast, current pain intensity has consistently been one of most successful predictors of future pain (Andersson, 2004; Gandhi et al., 2020; Landmark et al., 2019; Yarnitsky et al., 2008). This investigation improves upon this previous work by explaining the equivocal nature of QST in predicting outcome: risk for worse pain outcome is possibly not due to alterations in normal interpretation of specific nociceptive modalities, but rather MMH. When testing whether current pain (baseline pelvic pain), MMH, or nociceptive modality-specific components best predicted future pelvic pain outcome, we found that current pain provided a diminishing degree of predictive power annually, while MMH provided an increasing degree of predictive power over time. Additionally, MMH was the only significant predictor of year four pelvic pain intensity. Factors underlying modality-specific effects of QST did not predict outcome at any year other than baseline (see Slade et al., 2014), except for a modest correlation between baseline bladder hypersensitivity and pelvic pain. Therefore, the modality-specific QST differences often observed in cross-sectional studies comparing pain patients to controls (e.g., Greenspan et al., 2020; Harte et al., 2019) could reflect dynamic states of hypersensitivity. In contrast, the increased MMH in participants that develop worse pelvic pain may indicate that a pervasive sensory processing mechanism is persistently involved.

### 4.3 Strengths and Limitations

There are limitations that may affect the generalizability of our results. The sample included a large number of college students from neighboring universities. As a result, the sample was young (*M* =25, *SD* =6 years) and the vast majority were nulliparous (85% without a prior pregnancy). Our multimodal QST panel requires extensive training of personnel and is time consuming for the participant (*>* 3 hours including self-report questionnaires); therefore, it may be less feasible to administer in a clinical setting. Because the baseline QST assessment was conducted on a single day, and the stability of MMH is unknown, MMH may vary day to day. With that in mind, it is remarkable that MMH derived from a one-day QST panel predicted average pelvic pain in the prior week four years into the future.

Despite these limitations, the multimodal QST panel used here is one of the largest efforts to characterize MMH in individuals harboring variable degrees of risk for CPP and includes one of the longest follow-up periods of any previous QST pelvic pain study. The current investigation leveraged PCA that facilitated dimensionality reduction (from 40 to 3 variables) in a racially diverse sample enriched with at-risk individuals.

## 5 Conclusion

This analysis provided crucial experimental evidence of MMH, a hypothesized construct underlying “centralized” mechanisms of pain sensitivity, and its predictive ability of worse (pelvic) pain outcome. Our study demonstrates that focusing on MMH rather than single modalities can improve the prediction of pain status or outcome. Moreover, medical and psychological professional organizations have been reluctant to adopt diagnostics for sensory sensitivity (e.g., sensory processing disorder; Byrne, 2009; Section On Complementary And Integrative Medicine et al., 2012). Conversely, our results challenge this denial by showing that sensory sensitivity is more than a yes/no diagnosis. MMH exists on a continuum, and hypersensitive individuals are more vulnerable to worse future pain (Bar-Shalita et al., 2019). Neuroimaging paradigms have implicated the anterior insula and cingulate cortex as important for multimodal sensory integration and nociceptive appraisal (Harte et al., 2016; López-Solá et al., 2014). Therefore, future work focused on reversing the course of chronic pain would be well served to understand and target the neural mechanisms that underlie MMH.

## Data Availability

All code and data for this investigation is available on GitHub (https://github.com/mkmiecik14/mmh) and OSF (link will be inserted here upon publication).

## 6 Acknowledgments

The authors thank Dr. GF Gebhart for advice on study deign, interpretation, and editorial assistance. The authors are also grateful for our lab staff performing the QST assessments and participants for volunteering their time. This study was funded by the National Institute of Child and Human Development (R01HD098193) and the National Institute of Diabetes and Digestive and Kidney Diseases (R01DK100368).

## Notes

### Competing Interest Statement

The authors have declared no competing interest.

### Author Declarations

The Institutional Review Board of NorthShore University HealthSystem gave ethical approval for this work.

## References

Abdi, H. (2007). Singular value decomposition (SVD) and generalized singular value decomposition (GSVD). Sage.

Abdi, H., & Williams, L. J. (2010). Principal component analysis. Wiley Interdisciplinary Reviews: Computational Statistics, 2 (4), 433–459. https://doi.org/10.1002/wics.101

Andersson, H. (2004). The course of non-malignant chronic pain: A 12-year follow-up of a cohort from the general population. European Journal of Pain, 8 (1), 47–53. https://doi.org/10.1016/S1090-3801(03)00064-8

Arendt-Nielsen, L. (2017). Mechanistic similarities between fibromyalgia and other chronic pain conditions. PAIN Reports, 2 (1), 3. https://doi.org/ https://doi.org/10.1097/PR9.0000000000000582

Aykan, S., Vatansever, G., Doğanay-Erdoğan, B., & Kalaycıoğlu, C. (2020). Development of sensory sensitivity scales (SeSS): Reliability and validity analyses. Research in Developmental Disabilities, 100, 103612. https://doi.org/10.1016/j.ridd.2020.103612

Bar-Shalita, T., Granovsky, Y., Parush, S., & Weissman-Fogel, I. (2019). Sensory modulation disorder (SMD) and pain: A new perspective. Frontiers in Integrative Neuroscience, 13, 27. https://doi.org/10.3389/fnint.2019.00027

Beaton, D., Chin Fatt, C. R., & Abdi, H. (2014). An ExPosition of multivariate analysis with the singular value decomposition in r. Computational Statistics & Data Analysis, 72, 176–189. https://doi.org/10.1016/j.csda.2013.11.006

Berry, S. H., Elliott, M. N., Suttorp, M., Bogart, L. M., Stoto, M. A., Eggers, P., Nyberg, L., & Clemens, J. Q. (2011). Prevalence of symptoms of bladder pain syndrome/interstitial cystitis among adult females in the united states. Journal of Urology, 186 (2), 540–544. https://doi.org/10.1016/j.juro.2011.03.132

Bhalang, K., Sigurdsson, A., Slade, G. D., & Maixner, W. (2005). Associations among four modalities of experimental pain in women. The Journal of Pain, 6 (9), 604–611. https://doi.org/10.1016/j.jpain.2005.04.006

Birket-Smith, M. (2001). Somatization and chronic pain. Acta Anaesthesiologica Scandinavica, 45 (9), 1114–1120. https://doi.org/10.1034/j.1399-6576.2001.450911.x

Bordeleau, M., Barron, D., Léonard, G., & Backonja, M. (2021). Realigning the role of quantitative sensory testing in sensory profiling of patients with and without neuropathic pain. PAIN, 162 (11), 2780. https://doi.org/10.1097/j.pain.0000000000002378

Brown, C., Tollefson, N., Dunn, W., Cromwell, R., & Filion, D. (2001). The adult sensory profile: Measuring patterns of sensory processing. The American Journal of Occupational Therapy, 55 (1), 75–82. https://doi.org/10.5014/ajot.55.1.75

Byrne, M. W. (2009). Sensory processing disorder: Any of a nurse practitioner’s business? Journal of the American Academy of Nurse Practitioners, 21 (6), 314–321. https://doi.org/10.1111/j.1745-7599.2009.00417.x

Cathcart, S., Winefield, A. H., Rolan, P., & Lushington, K. (2009). Reliability of temporal summation and diffuse noxious inhibitory control [Publisher: Hindawi]. Pain Research and Management, 14 (6), 433–438. https://doi.org/10.1155/2009/523098

Cruz-Almeida, Y., & Fillingim, R. B. (2014). Can quantitative sensory testing move us closer to mechanism-based pain management? Pain Medicine, 15 (1), 61–72. https://doi.org/10.1111/pme.12230

Curatolo, M. (2011). Diagnosis of altered central pain processing. Spine, 36, S200. https://doi.org/10.1097/BRS.0b013e3182387f3d

Curatolo, M., Arendt-Nielsen, L., & Petersen-Felix, S. (2006). Central hypersensitivity in chronic pain: Mechanisms and clinical implications. Physical Medicine and Rehabilitation Clinics of North America, 17 (2), 287–302. https://doi.org/10.1016/j.pmr.2005.12.010

Dahlhamer, J., Lucas, J., Zelaya, C., Nahin, R., Mackey, S., DeBar, L., Kerns, R., Von Korff, M., Porter, L., & Helmick, C. (2018). Prevalence of chronic pain and high-impact chronic pain among adults — united states, 2016. MMWR. Morbidity and Mortality Weekly Report, 67 (36), 1001–1006. https://doi.org/10.15585/mmwr.mm6736a2

Fernandes, C., Pidal-Miranda, M., Samartin-Veiga, N., & Carrillo-de-la-Peña, M. T. (2019). Conditioned pain modulation as a biomarker of chronic pain: A systematic review of its concurrent validity. PAIN, 160 (12), 2679–2690. https://doi.org/10.1097/j.pain.0000000000001664

Fitzcharles, M.-A., Cohen, S. P., Clauw, D. J., Littlejohn, G., Usui, C., & Häuser, W. (2021). Nociplastic pain: Towards an understanding of prevalent pain conditions. The Lancet, 397 (10289), 2098–2110. https://doi.org/10.1016/S0140-6736(21)00392-5

FitzGerald, M. P., Koch, D., & Senka, J. (2005). Visceral and cutaneous sensory testing in patients with painful bladder syndrome. Neurourology and Urodynamics, 24 (7), 627–632. https://doi.org/10.1002/nau.20178

Forstenpointner, J., Ruscheweyh, R., Attal, N., Baron, R., Bouhassira, D., Enax-Krumova, E. K., Finnerup, N. B., Freynhagen, R., Gierthmühlen, J., Hansson, P., Jensen, T. S., Maier, C., Rice, A. S. C., Segerdahl, M., Tölle, T., Treede, R.-D., & Vollert, J. (2021). No pain, still gain (of function): The relation between sensory profiles and the presence or absence of self-reported pain in a large multicenter cohort of patients with neuropathy. PAIN, 162 (3), 718–727. https://doi.org/10.1097/j.pain.0000000000002058

Gandhi, W., Pomares, F. B., Naso, L., Asenjo, J.-F., & Schweinhardt, P. (2020). Neuropathic pain after thoracotomy: Tracking signs and symptoms before and at monthly intervals following surgery. European Journal of Pain, 24 (7), 1269–1289. https://doi.org/10.1002/ejp.1569

Geisser, M. E., Glass, J. M., Rajcevska, L. D., Clauw, D. J., Williams, D. A., Kileny, P. R., & Gracely, R. H. (2008). A psychophysical study of auditory and pressure sensitivity in patients with fibromyalgia and healthy controls. The Journal of Pain, 9 (5), 417–422. https://doi.org/10.1016/j.jpain.2007.12.006

Georgopoulos, V., Akin-Akinyosoye, K., Zhang, W., McWilliams, D. F., Hendrick, P., & Walsh, D. A. (2019). Quantitative sensory testing and predicting outcomes for musculoskeletal pain, disability, and negative affect: A systematic review and meta-analysis. Pain, 160 (9), 1920–1932. https://doi.org/10.1097/j.pain.0000000000001590

Greenspan, J. D., Slade, G. D., Bair, E., Dubner, R., Fillingim, R. B., Ohrbach, R., Knott, C., Diatchenko, L., Liu, Q., & Maixner, W. (2013). Pain sensitivity and autonomic factors associated with development of TMD: The OPPERA prospective cohort study. The journal of pain : official journal of the American Pain Society, 14 (12), T63–74.e1–6. https://doi.org/10.1016/j.jpain.2013.06.007

Greenspan, J. D., Slade, G. D., Bair, E., Dubner, R., Fillingim, R. B., Ohrbach, R., Knott, C., Mulkey, F., Rothwell, R., & Maixner, W. (2011). Pain sensitivity risk factors for chronic TMD: Descriptive data and empirically identified domains from the OPPERA case control study. The Journal of Pain, 12 (11), T61–T74. https://doi.org/10.1016/j.jpain.2011.08.006

Greenspan, J. D., Slade, G. D., Rathnayaka, N., Fillingim, R. B., Ohrbach, R., & Maixner, W. (2020). Experimental pain sensitivity in subjects with temporomandibular disorders and multiple other chronic pain conditions: The OPPERA prospective cohort study. Journal of Oral & Facial Pain and Headache, 34, s43–s56. https://doi.org/10.11607/ofph.2583

Grundström, H., Larsson, B., Arendt-Nielsen, L., Gerdle, B., & Kjølhede, P. (2019). Associations between pain thresholds for heat, cold and pressure, and pain sensitivity questionnaire scores in healthy women and in women with persistent pelvic pain. European Journal of Pain, 23 (9), 1631–1639. https://doi.org/10.1002/ejp.1439

Hanno, P. M., Burks, D. A., Clemens, J. Q., Dmochowski, R. R., Erickson, D., FitzGerald, M. P., Forrest, J. B., Gordon, B., Gray, M., Mayer, R. D., Newman, D., Nyberg, L., Payne, C. K., Wesselmann, U., & Faraday, M. M. (2011). AUA guideline for the diagnosis and treatment of interstitial cystitis/bladder pain syndrome. Journal of Urology, 185 (6), 2162–2170. https://doi.org/10.1016/j.juro.2011.03.064

Harper, D., Schrepf, A., & Clauw, D. (2016). Pain mechanisms and centralized pain in temporomandibular disorders. Journal of Dental Research, 95 (10), 1102–1108. https://doi.org/10.1177/0022034516657070

Harte, S. E., Ichesco, E., Hampson, J. P., Peltier, S. J., Schmidt-Wilcke, T., Clauw, D. J., & Harris, R. E. (2016). Pharmacologic attenuation of cross-modal sensory augmentation within the chronic pain insula: PAIN, 157 (9), 1933–1945. https://doi.org/10.1097/j.pain.0000000000000593

Harte, S. E., Schrepf, A., Gallop, R., Kruger, G. H., Lai, H. H. H., Sutcliffe, S., Halvorson, M., Ichesco, E., Naliboff, B. D., Afari, N., Harris, R. E., Farrar, J. T., Tu, F., Landis, J. R., Clauw, D. J., & for the MAPP Research Network. (2019). Quantitative assessment of nonpelvic pressure pain sensitivity in urologic chronic pelvic pain syndrome: A MAPP research network study. Pain, 160 (6), 1270–1280. https://doi.org/10.1097/j.pain.0000000000001505

Hastie, B. A., Riley, J. L., Robinson, M. E., Glover, T., Campbell, C. M., Staud, R., & Fillingim, R. B. (2005). Cluster analysis of multiple experimental pain modalities. Pain, 116 (3), 227–237. https://doi.org/10.1016/j.pain.2005.04.016

Hellman, K. M., Datta, A., Steiner, N. D., Kane Morlock, J. N., Garrison, E. F., Clauw, D. J., & Tu, F. F. (2018). Identification of experimental bladder sensitivity among dysmenorrhea sufferers. American Journal of Obstetrics and Gynecology, 219 (1), 84.e1–84.e8. https://doi.org/10.1016/j.ajog.2018.04.030

Hellman, K. M., Patanwala, I. Y., Pozolo, K. E., & Tu, F. F. (2015). Multimodal nociceptive mechanisms underlying chronic pelvic pain. American Journal of Obstetrics and Gynecology, 213 (6), 827.e1–827.e9. https://doi.org/10.1016/j.ajog.2015.08.038

Hellman, K. M., Roth, G. E., Dillane, K. E., Garrison, E. F., Oladosu, F. A., Clauw, D. J., & Tu, F. F. (2020). Dysmenorrhea subtypes exhibit differential quantitative sensory assessment profiles. PAIN, 161 (6), 1227– 1236. https://doi.org/10.1097/j.pain.0000000000001826

Hollins, M., Harper, D., Gallagher, S., Owings, E. W., Lim, P. F., Miller, V., Siddiqi, M. Q., & Maixner, W. (2009). Perceived intensity and unpleasantness of cutaneous and auditory stimuli: An evaluation of the generalized hypervigilance hypothesis: Pain, 141 (3), 215–221. https://doi.org/10.1016/j.pain.2008.10.003

Janal, M. N., Glusman, M., Kuhl, J. P., & Clark, W. C. (1994). On the absence of correlation between responses to noxious heat, cold, electrical and ischemie stimulation. Pain, 58 (3), 403–411. https://doi.org/10.1016/0304-3959(94)90135-X

Jensen, M. P., Wang, W., Potts, S. L., & Gould, E. M. (2012). Reliability and validity of individual and composite recall pain measures in patients with cancer. Pain Medicine, 13 (10), 1284–1291. https://doi.org/10.1111/j.1526-4637.2012.01470.x

Kaya, S., Hermans, L., Willems, T., Roussel, N., & Meeus, M. (2013). Central sensitization in urogynecological chronic pelvic pain : A systematic literature review [Number: 4]. PAIN PHYSICIAN, 16 (4), 291–308. Retrieved January 21, 2022, from http://hdl.handle.net/1854/LU-4109235

Kmiecik, M. J., Tu, F. F., Silton, R. L., Dillane, K. E., Roth, G. E., Harte, S. E., & Hellman, K. M. (2021). Cortical mechanisms of visual hypersensitivity in women at risk for chronic pelvic pain. PAIN. https://doi.org/10.1097/j.pain.0000000000002469

Landmark, T., Romundstad, P., Butler, S., Kaasa, S., & Borchgrevink, P. (2019). Development and course of chronic widespread pain: The role of time and pain characteristics (the HUNT pain study). PAIN, 160 (9), 1976– 1981. https://doi.org/10.1097/j.pain.0000000000001585

Lautenbacher, S., & Rollman, G. B. (1993). Sex differences in responsiveness to painful and non-painful stimuli are dependent upon the stimulation method. Pain, 53 (3), 255–264. https://doi.org/10.1016/0304-3959(93)90221-A

Li, R., Li, B., Kreher, D. A., Benjamin, A. R., Gubbels, A., & Smith, S. M. (2020). Association between dysmenorrhea and chronic pain: A systematic review and meta-analysis of population-based studies. American Journal of Obstetrics and Gynecology, 223 (3), 350–371. https://doi.org/10.1016/j.ajog.2020.03.002

Lionetti, F., Aron, A., Aron, E. N., Burns, G. L., Jagiellowicz, J., & Pluess, M. (2018). Dandelions, tulips and orchids: Evidence for the existence of low-sensitive, medium-sensitive and high-sensitive individuals. Translational Psychiatry, 8 (1), 24. https://doi.org/10.1038/s41398-017-0090-6

López-Solá, M., Pujol, J., Wager, T. D., Garcia-Fontanals, A., Blanco-Hinojo, L., Garcia-Blanco, S., Poca-Dias, V., Harrison, B. J., Contreras-Rodríguez, O., Monfort, J., Garcia-Fructuoso, F., & Deus, J. (2014). Altered functional magnetic resonance imaging responses to nonpainful sensory stimulation in fibromyalgia patients: Brain response to nonpainful multi-sensory stimulation in fibromyalgia. Arthritis & Rheumatology, 66 (11), 3200–3209. https://doi.org/10.1002/art.38781

Martenson, M. E., Halawa, O. I., Tonsfeldt, K. J., Maxwell, C. A., Hammack, N., Mist, S. D., Pennesi, M. E., Bennett, R. M., Mauer, K. M., Jones, K. D., & Heinricher, M. M. (2016). A possible neural mechanism for photosensitivity in chronic pain. PAIN, 157 (4), 868–878. https://doi.org/10.1097/j.pain.0000000000000450

Morris, M. C., Bruehl, S., Stone, A. L., Garber, J., Smith, C., Palermo, T. M., & Walker, L. S. (2021). Does quantitative sensory testing improve prediction of chronic pain trajectories? a longitudinal study of youth with functional abdominal pain participating in a randomized controlled trial of cognitive behavioral treatment. The Clinical Journal of Pain, 37 (9), 648–656. https://doi.org/10.1097/AJP.0000000000000956

Müller, M., Bütikofer, L., Andersen, O. K., Heini, P., Arendt-Nielsen, L., Jüni, P., & Curatolo, M. (2021). Cold pain hypersensitivity predicts trajectories of pain and disability after low back surgery: A prospective cohort study. Pain, 162 (1), 184–194. https://doi.org/10.1097/j.pain.0000000000002006

Myles, P. S., Troedel, S., Boquest, M., & Reeves, M. (1999). The pain visual analog scale: Is it linear or nonlinear? Anesthesia & Analgesia, 89 (6), 1517. https://doi.org/10.1213/00000539-199912000-00038

Nir, R.-R., & Yarnitsky, D. (2015). Conditioned pain modulation. Current Opinion in Supportive and Palliative Care, 9 (2), 131–137. https://doi.org/10.1097/SPC.0000000000000126

O’Brien, A. T., Deitos, A., Triñanes Pego, Y., Fregni, F., & Carrillo-de-la-Peña, M. T. (2018). Defective endogenous pain modulation in fibromyalgia: A meta-analysis of temporal summation and conditioned pain modulation paradigms. The Journal of Pain, 19 (8), 819–836. https://doi.org/10.1016/j.jpain.2018.01.010

Rolke, R., Baron, R., Maier, C., Tölle, T. R., Treede, R..-., Beyer, A., Binder, A., Birbaumer, N., Birklein, F., Bötefür, I. C., Braune, S., Flor, H., Huge, V., Klug, R., Landwehrmeyer, G. B., Magerl, W., Maihöfner, C., Rolko, C., Schaub, C., … Wasserka, B. (2006). Quantitative sensory testing in the german research network on neuropathic pain (DFNS): Standardized protocol and reference values. PAIN, 123 (3), 231–243. https://doi.org/10.1016/j.pain.2006.01.041

Sandri, A., Cecchini, M. P., Riello, M., Zanini, A., Nocini, R., Fiorio, M., & Tinazzi, M. (2021). Pain, smell, and taste in adults: A narrative review of multisensory perception and interaction. Pain and Therapy. https://doi.org/10.1007/s40122-021-00247-y

Santana, A. N., de Santana, C. N., & Montoya, P. (2020). Chronic pain diagnosis using machine learning, questionnaires, and QST: A sensitivity experiment. Diagnostics, 10 (11), 958. https://doi.org/10.3390/diagnostics10110958

Schmelz, M. (2020). What can we learn from the failure of quantitative sensory testing? PAIN, 162 (3), 663–664. https://doi.org/10.1097/j.pain.0000000000002059

Schoep, M. E., Nieboer, T. E., van der Zanden, M., Braat, D. D., & Nap, A. W. (2019). The impact of menstrual symptoms on everyday life: A survey among 42,879 women. American Journal of Obstetrics and Gynecology, 220 (6), 569.e1–569.e7. https://doi.org/10.1016/j.ajog.2019.02.048

Schrepf, A., Williams, D. A., Gallop, R., Naliboff, B. D., Basu, N., Kaplan, C., Harper, D. E., Landis, J. R., Clemens, J. Q., Strachan, E., Griffith, J. W., Afari, N., Hassett, A., Pontari, M. A., Clauw, D. J., & Harte, S. E. (2018). Sensory sensitivity and symptom severity represent unique dimensions of chronic pain: A MAPP research network study. PAIN, 159 (10), 2002–2011. https://doi.org/10.1097/j.pain.0000000000001299

Section On Complementary and Integrative Medicine, on Children with Disabilities, C., Zimmer, M., Desch, L., Rosen, L. D., Bailey, M. L., Becker, D., Culbert, T. P., McClafferty, H., Sahler, O. J. Z., Vohra, S., Liptak, G. S., Adams, R. C., Burke, R. T., Friedman, S. L., Houtrow, A. J., Kalichman, M. A., Kuo, D. Z., Levy, S. E., … Wiley, S. E. (2012). Sensory integration therapies for children with developmental and behavioral disorders. Pediatrics, 129 (6), 1186–1189. https://doi.org/10.1542/peds.2012-0876

Slade, G. D., Sanders, A. E., Ohrbach, R., Fillingim, R. B., Dubner, R., Gracely, R. H., Bair, E., Maixner, W., & Greenspan, J. D. (2014). Pressure pain thresholds fluctuate with - but do not usefully predict - the clinical course of painful temporomandibular disorder. Pain, 155 (10), 2134– 2143. https://doi.org/10.1016/j.pain.2014.08.007

Sriwatanakul, K., Kelvie, W., Lasagna, L., Calimlim, J. F., Weis, O. F., & Mehta, G. (1983). Studies with different types of visual analog scales for measurement of pain. Clinical Pharmacology & Therapeutics, 34 (2), 234–239. https://doi.org/10.1038/clpt.1983.159

Treede, R.-D. (2019). The role of quantitative sensory testing in the prediction of chronic pain. Pain, 160 (1), S66–S69. https://doi.org/10.1097/j.pain.0000000000001544

Tu, F. F., Epstein, A. E., Pozolo, K. E., Sexton, D. L., Melnyk, A. I., & Hellman, K. M. (2013). A noninvasive bladder sensory test supports a role for dysmenorrhea increasing bladder noxious mechanosensitivity. The Clinical Journal of Pain, 29 (10), 883–890. https://doi.org/10.1097/AJP.0b013e31827a71a3

Tu, F. F., Kane, J. N., & Hellman, K. M. (2017). Noninvasive experimental bladder pain assessment in painful bladder syndrome. BJOG: An International Journal of Obstetrics & Gynaecology, 124 (2), 283–291. https://doi.org/10.1111/1471-0528.14433

Wilbarger, J. L., & Cook, D. B. (2011). Multisensory hypersensitivity in women with fibromyalgia: Implications for well being and intervention. Archives of Physical Medicine and Rehabilitation, 92 (4), 653–656. https://doi.org/10.1016/j.apmr.2010.10.029

Yarnitsky, D., Crispel, Y., Eisenberg, E., Granovsky, Y., Ben-Nun, A., Sprecher, E., Best, L.-A., & Granot, M. (2008). Prediction of chronic post-operative pain: Pre-operative DNIC testing identifies patients at risk. Pain, 138 (1), 22–28. https://doi.org/10.1016/j.pain.2007.10.033

Yarnitsky, D., Granot, M., Nahman-Averbuch, H., Khamaisi, M., & Granovsky, Y. (2012). Conditioned pain modulation predicts duloxetine efficacy in painful diabetic neuropathy. Pain, 153 (6), 1193–1198. https://doi.org/10.1016/j.pain.2012.02.021

Zondervan, K. T., Yudkin, P. L., Vessey, M. P., Jenkinson, C. P., Dawes, M. G., Barlow, D. H., & Kennedy, S. H. (2001). Chronic pelvic pain in the community—symptoms, investigations, and diagnoses. American Journal of Obstetrics and Gynecology, 184 (6), 1149–1155. https://doi.org/10.1067/mob.2001.112904

