## Supplemental Material for "Multimodal Hypersensitivity Derived from Quantitative Sensory Testing Predicts Long-Term Pelvic Pain Outcome"

This document contains supplementary material referenced in the main body of the manuscript.

### 1 Methods

#### 1.1 Participants

Participants were recruited by public advertising and flyers in Evanston, Illinois and the surrounding communities. Severity of menstrual pain was confirmed with internet-based prospective symptom diaries for 1-2 months prior to full enrollment (Hellman et al., 2020). Participants were excluded for the presence of active pelvic or abdominal malignancies, absence of regular menses (except the chronic pain without BPS group), active genitourinary infection in the last four weeks, inability to read or comprehend the informed consent in English, refusal to undergo pelvic examination/testing, hypertension, or refusal to withdraw from oral contraceptives for two months prior to the study visit.

From the 354 participants, $n$=154 were excluded for the following reasons: 52 participants were missing one or more data points from QST (e.g., declined participation in task(s), equipment malfunction, migraine sensitivity precluded participation in visual stimulation, etc.) and one participant was under the influence of recreational or illicit substances during the testing appointment. Additionally, 25 participants were lost to follow-up, 16 participants withdrew from the study, and 60 participants were disqualified (e.g., over-recruited dysmenorrhea without bladder pain, started oral contraceptives, unable to complete protocol).

From the 200 women with complete QST data, 22 were randomized into a 12-month clinical trial that evaluated the efficacy of cyclical ($n$=4) and continuous ($n$=12) oral contraceptive pills (OCPs) of treating menstrual and bladder pain compared to a control group that did not take OCPs ($n$=6). Because of the low adherence to OCP use and small comparative sample sizes, we did not consider participants' enrollment in the clinical trial as exclusionary from our analyses of subsequent annual questionnaires. In addition, QST was performed on all participants before beginning use of OCPS and so data on internal relationships of MMH of longitudinal outcome was not expected to be confounded by OCP use.

Annual questionnaires were a reduced version of what was asked at their screen and baseline assessment visit and were used to assess pelvic pain outcome.

#### 1.2 Measures

##### 1.2.1 Quantitative Sensory Testing (QST)

**Bladder Filling Test** We developed a non-invasive bladder filling task (see Tu et al., 2013) to better characterize visceral hypersensitivity that is observed across CPP conditions like bladder pain syndrome and irritable bowel syndrome (Aslam et al., 2009; FitzGerald et al., 2005; Kanazawa et al., 2011). This task has been validated for identifying abnormalities associated with bladder pain syndrome, chronic pelvic pain and even small changes associated with increased daily bladder symptoms (Hellman et al., 2018; Tu et al., 2017). After voiding their bladder, participants ingested 20 fluid ounces of water and rated their bladder pain and urgency on a 0-100 VAS across four time points: baseline, first sensation (FS) of bladder filling, first urge (FU) which is the normal desire to void their bladder, and maximum tolerance (MT) of bladder filling (corresponding to widely used cystometric sensory thresholds Haylen et al., 2010). After reaching maximum tolerance (or 2 hours) and voiding their bladder, participants rated their perceived bladder pain on 0-100 VAS according to four McGill pain questionnaire descriptors to potentially differentiate A$\delta$ from C fiber pain components: sharp, pressing, dull, and prickling (Beissner et al., 2010). In sum, a total of 12 measures from the bladder task were included in this analysis (i.e., 4 provoked bladder pain ratings and 4 bladder urgency ratings across the time points, and 4 McGill descriptor ratings at completion).

**Pressure Pain Thresholds (PPTs)** We examined PPTs both transvaginally and externally because local alterations in myofascial pelvic sensitivity (Hellman et al., 2015) and widespread alterations in bodily sensitivity are thought to underlie centralized pain (Granges & Littlejohn, 1993) that may contribute to chronic pelvic pain. We determined participants' PPTs using a digital algometer with a 1-cm$^2$ rubber tip driven at a ramp rate of 4 Newtons(N)/sec at three fibromyalgia tender point sites (Wolfe et al., 1990)—right trapezius, the right medial knee fat pad, and the right greater trochanter (hip)—and the forehead. Vaginal PPTs were measured using a finger mounted 1-cm$^2$ diameter force-sensing resistor at a ramp rate of 0.5 N/sec at four vaginal sites: right (5 o'clock position) and left iliococcygeus (7 o'clock), anteriorly against the bladder (12 o'clock), and posteriorly against the anorectal raphe (6 o'clock). Body and vaginal PPT procedures utilized software that guided stimulus application and resulted in high ($\alpha > .89$) inter- and intra-examiner reliability (for details see Hellman et al., 2020). After each set of PPTs, participants were asked to rate their pain on a 0-10 NRS at each site. These after-pain ratings were adjusted for baseline pain ratings recorded before PPT procedures. We previously established that PPTs and after-pain represent two different components of sensation contributing to MMH (Hellman et al., 2015). PPTs represent the average force from two independent trials separated by 2 a minute break period. These averaged PPTs were multiplied by -1 so that a greater value indicates increased sensitivity. In total, 16 PPT measures were included in the PCA: 8 PPTs (4 body and 4 vaginal sites) and 8 after-pain ratings (4 body and 4 vaginal sites).

**Conditioned Pain Modulation (CPM)** We included CPM in our QST panel because prior studies have demonstrated that CPM predicts pain outcome (Yarnitsky et al., 2008; Yarnitsky et al., 2012). CPM efficiency is thought to be a metric of descending inhibition as tested by "pain inhibits pain" paradigms (Nir & Yarnitsky, 2015). We assessed participants' CPM by repeat PPT testing of the left medial knee fat pad before and after ice water immersion of the contralateral hand. After an initial PPT measurement, participants waited two-minutes before submerging their right hand up to their wrist into a bath of circulating water maintained at 0-6°C. After 10 seconds of submersion, participants rated their hand/cold pain on a 0-10 NRS and after 20 seconds of submersion, a repeat PPT measure was taken from the left medial knee fat pad, after which participants were allowed to remove their hand from the cold water bath. CPM was calculated by subtracting the PPT force (in Newtons) taken before the water bath from the PPT taken after the water bath (i.e., CPM = After-Before). CPM values were then multiplied by -1 so that a greater number denoted reduced/inefficient CPM (i.e., increased impairment) before including results in the PCA. Additionally, cold pain ratings were adjusted for the water temperature by extracting the residuals from a linear model predicting cold pain as a function of water temperature. Therefore, a total of 2 measures from CPM testing were included into the PCA: 1 CPM score and 1 cold pain rating adjusted for water temperature.

**Temporal Summation (TS)** Increased response to repeated application of noxious stimuli (i.e., wind-up pain) is thought to reflect a unique component of spinally mediated sensitization that may be associated with increased risk of chronic pain (Cathcart et al., 2009; O'Brien et al., 2018; Thompson et al., 2020). Therefore, we measured TS using a commonly used strategy: 10 pressure pulses delivered to the right medial knee fat pad using the same body PPT algometer as described above (Cathcart et al., 2009). Each pulse was delivered at a ramp rate of 4 N/sec with 1 second breaks between pulses using a software-based metronome to guide application. Each pulse was applied until the initial threshold for a pain rating of 1 was reached. Participants rated their baseline pain at the application site on a 0-10 NRS following each pulse. The TS task ended when participants reported a pain rating of 6 or after the tenth trial. Each participant's baseline pain was subtracted from her pain ratings collected after each pulse. A total of three TS measures were entered into the PCA: the average pain experienced during TS, the rate of change in pain ratings as a function of trial (i.e., slope), and the maximum TS trial experienced. The maximum TS trial was multiplied by -1 so that greater values indicated increased sensitivity (i.e., fewer trials allowed due to reaching a pain rating of 6).

**Visual Stimulation** Investigations of MMH are well served to include additional sensory modalities, such as vision, given that light hypersensitivity is commonly reported in conditions with generalized sensory hypersensitivity, like fibromyalgia and migraine (Friedman & De Ver Dye, 2009; Harte et al., 2016; Martenson et al., 2016). We assessed participants' visual unpleasantness sensitivity by presenting a periodic pattern-reversal blue/yellow checkerboard stimulus alternating at 25 Hz for 20 seconds across five blocks. Each block contained a single maximal brightness intensity (1, 30, 60, 90, or 120 lux) and block order was randomized across participants. After each block, participants rated stimulus unpleasantness using the Gracely Box Scale that lists the numbers 0 to 20 next to a set of verbal anchors (Gracely & Kwilosz, 1988). A total of two visual sensitivity measures were entered into the PCA: the average visual unpleasantness rating across the blocks, and the rate of change in visual unpleasantness as a function of brightness intensity (Kmiecik et al., 2021).

**Auditory Stimulation** Likewise, auditory stimulation provided an additional sensory modality with reported hypersensitivities in functional pain syndromes (Hollins et al., 2009; López-Solà et al., 2014; Wilbarger & Cook, 2011). Prior to assessing the participants' auditory unpleasantness sensitivity, an audio program first verified that participants maintained less than 20 dB hearing loss (250-8000 Hz) (Picou et al., 2013) to equate hearing ability. Next, we presented a series of auditory steady state 80Hz (D'haenens et al., 2009) volume modulated tones (1200 and 1350Hz) in random order of intensity (15, 30 45, or 60 dB) delivered via ground-isolated optimally flat frequency response pneumatic insert earphones (Etymotic, Elk Grove Village, IL). After each 20 second stimulus, the participant rated her perceived unpleasantness on the Gracely Box Scale as described above. A total of two auditory sensitivity measures were entered into the PCA: the average auditory unpleasantness rating across the blocks, and the rate of change in auditory unpleasantness as a function of loudness intensity (i.e., slope).

We recorded participants' scalp electroencephalography (EEG) during both visual and auditory sensitivity tasks. Given the behavioral focus of this investigation, these EEG data were out of scope and not included in the present investigation. EEG data from the visual task are published elsewhere (Kmiecik et al., 2021).

##### 1.2.2 Self-Report Questionnaires

As part of data collection on the participants' health history, we administered several validated self-report questionnaires that assessed various aspects of pelvic pain and associated symptomologies. Bladder pain syndrome is a significant CPP condition (Hanno et al., 2011) and was assessed via the Interstitial Cystitis Symptom Inventory (ICSI) and IC Problem Index (ICPI) (O'Leary et al., 1997). The Genitourinary Pain Index (GUPI) provided complementary assessment of these urogenital symptoms (Clemens et al., 2009). The Complex Medical Symptoms Inventory (CMSI) was used to assess symptoms indicative of fibromyalgia (Williams & Schilling, 2009) and included assessments over the lifetime and last year. Somatic symptoms were assessed using the Brief Symptom Inventory (Derogatis & Melisaratos, 1983). We assessed several health domains from the NIH Patient Reported Outcomes Measurement System (PROMIS; Broderick et al., 2013), including anxiety, depression, pain interference, pain behavior, global physical health, and global mental health.

Also, menstrual pain is associated with non-cyclic pelvic pain (Westling et al., 2013) and non-pelvic pain hypersensitivity (Hellman et al., 2020; Iacovides et al., 2015; Payne et al., 2017; Payne et al., 2019), suggesting that menstrual pain may be a risk factor for developing chronic pain (Iacovides et al., 2015; Payne et al., 2017). Therefore, we assessed participants' menstrual pain on a 0-100 VAS on the worst day of their period over the past three months in the absence of pain relievers (e.g., non-steroidal anti-inflammatory drugs [NSAIDs], acetaminophen, etc.).

#### 1.3 Statistical Analyses

Permutation testing for the PCA was conducted by creating null distributions for each principal component (PC) by shuffling each column's values without replacement and repeating the PCA for 2,000 iterations. Probability values for each PC were calculated by comparing our fixed-effects eigenvalues to their respective null distributions. Bootstrapping assessed the stability of the measures and the extent to which they significantly contributed to the variance of a PC. Bootstrap samples were formed by selecting participants at random with replacement. Bootstrap distributions were formed by supplementary projecting the bootstrap samples onto the eigen-space generated from the fixed-effects analysis (see Beaton et al., 2014). This procedure was repeated 2,000 times.

Analyses were performed in R (4.1.0) within RStudio (1.4.1106) using the following packages: fixed- and random-effects PCA was performed using *Ex-*

*Position* (Beaton et al., 2014), bootstrapped correlations were computed using *psych* (Revelle, 2020), effect sizes were calculated using *effectsize* (Ben-Shachar et al., 2020), data processing and figures were generated using *tidyverse* packages (Wickham et al., 2019), color palettes were inspired by *RColorBrewer* (Neuwirth, 2014) and *ghibli* (Henderson, 2020).

| QST Measure | *Mean* | *SD* | *Min* | *Max* |
|---|---|---|---|---|
| **PPT (Newtons)** | | | | |
| Vaginal 12 o'clock | 9.7 | 6.8 | 0.9 | 30.7 |
| Vaginal 5 o'clock | 9.0 | 5.6 | 0.4 | 31.0 |
| Vaginal 6 o'clock | 8.6 | 5.6 | 0.6 | 29.4 |
| Vaginal 7 o'clock | 7.2 | 4.9 | 0.8 | 30.1 |
| Forehead | 18.1 | 8.9 | 3.5 | 42.5 |
| Hip | 25.5 | 12.0 | 4.5 | 73.7 |
| Knee | 21.8 | 9.6 | 2.1 | 65.3 |
| Shoulder | 20.4 | 9.8 | 4.4 | 57.2 |
| **After-pain (0-10 NRS)** | | | | |
| Vaginal 12 o'clock | 1.4 | 1.0 | -4.0 | 6.0 |
| Vaginal 5 o'clock | 1.5 | 1.1 | -3.0 | 6.5 |
| Vaginal 6 o'clock | 1.4 | 1.0 | -3.0 | 7.0 |
| Vaginal 7 o'clock | 1.5 | 1.1 | -3.5 | 7.0 |
| Forehead | 1.2 | 0.6 | 0.0 | 5.0 |
| Hip | 1.3 | 0.7 | -0.5 | 5.5 |
| Knee | 1.2 | 0.6 | -0.5 | 3.5 |
| Shoulder | 1.2 | 0.6 | -3.0 | 4.5 |
| **Bladder Task (0-100 VAS)** | | | | |
| BL Pain | 5.1 | 10.7 | 0 | 57 |
| FS Pain | 8.4 | 13.4 | 0 | 66 |
| FU Pain | 16.2 | 19.9 | 0 | 100 |
| MT Pain | 30.7 | 29.5 | 0 | 93 |
| FS Urgency | 21.7 | 14.1 | 0 | 67 |
| FU Urgency | 48.8 | 16.2 | 2 | 100 |
| MT Urgency | 83.4 | 12.9 | 6 | 100 |
| **Bladder Descriptors (0-10 NRS)** | | | | |
| Dull | 27.3 | 28.1 | 0 | 90 |
| Pressing | 48.2 | 32.0 | 0 | 100 |
| Prickling | 13.9 | 20.6 | 0 | 87 |
| Sharp | 20.3 | 25.6 | 0 | 98 |
| **Pelvic Descriptors (0-10 NRS)** | | | | |
| Dull | 1.4 | 1.7 | 0 | 9 |
| Pressing | 3.3 | 2.3 | 0 | 10 |
| Prickling | 0.6 | 1.5 | 0 | 8 |
| Sharp | 2.0 | 1.9 | 0 | 8 |

Table 1: **Descriptive Statistics for PPTs and Bladder Task of CRAMPP Participants.** *Note.* After-pain ratings were adjusted by subtracting the baseline pain ratings prior to PPT testing. PPT=Pressure Pain Threshold; BL=Baseline; FS=First Sensation; FU=First Urge; MT=Maximum Tolerance; NRS=numeric rating scale; VAS=visual analog scale.

| QST Measure | *Mean* | *SD* | *Min* | *Max* |
|---|---|---|---|---|
| **Cold Pain (0-10 NRS)** | | | | |
| Rating | 5.49 | 2.34 | 0 | 10 |
| Residuals | -0.1 | 2.3 | -5.7 | 4.5 |
| **CPM** | | | | |
| Left Knee (Newtons) | 6.7 | 9.1 | -10.8 | 47.2 |
| **Temporal Summation** | | | | |
| Max Trial | 9.5 | 1.6 | 2 | 10 |
| Mean Pain (0-10 NRS) | 2.2 | 1.3 | -0.5 | 5.3 |
| Slope | 0.2 | 0.4 | -0.3 | 3.0 |
| **Visual Unpleasantness** | | | | |
| Mean (0-20 GBS) | 8.1 | 3.9 | 0.0 | 19.6 |
| Slope | 0.6 | 0.7 | -1.5 | 3.0 |
| **Auditory Unpleasantness** | | | | |
| Mean (0-20 GBS) | 6.4 | 2.5 | 1.2 | 13.6 |
| Slope | 1.9 | 0.8 | -1.3 | 4.0 |

Table 2: **Additional QST Descriptive Statistics (Cold Pain, CPM, TS, and Visual/Auditory Stimulation) of CRAMPP Participants.** *Note.* Cold pain residuals, not ratings, were used in the PCA. CPM=conditioned pain modulation; NRS=numeric rating scale; GBS=Gracely Box Scale.

| Year | Parameter | $b$ | $SE$ | $SS$ | $MSE$ | $F$ | $p$ |
|---|---|---|---|---|---|---|---|
| 1 | Intercept | 16.3 | 1.2 | 37298 | 194 | 191.9 | <.001 |
| | Baseline Pelvic Pain | 0.5 | 0.1 | 6641 | 194 | 34.2 | <.001 |
| | QST | 0.1 | 0.1 | 318 | 194 | 1.6 | 0.20 |
| | Bladder Test | 0.3 | 0.2 | 286 | 194 | 1.5 | 0.23 |
| | Audio/Visual | 0.2 | 0.5 | 37 | 194 | 0.2 | 0.66 |
| 2 | Intercept | 14.2 | 1.2 | 25407 | 186 | 135.8 | <.001 |
| | Baseline Pelvic Pain | 0.2 | 0.1 | 1171 | 186 | 6.3 | 0.01 |
| | QST | 0.1 | 0.1 | 209 | 186 | 1.1 | 0.29 |
| | Bladder Test | 0.4 | 0.2 | 546 | 186 | 2.9 | 0.09 |
| | Audio/Visual | 0.2 | 0.5 | 23 | 186 | 0.1 | 0.72 |
| 3 | Intercept | 12.8 | 1.3 | 15380 | 160 | 95.2 | <.001 |
| | Baseline Pelvic Pain | 0.2 | 0.1 | 907 | 160 | 5.6 | 0.02 |
| | QST | 0.2 | 0.1 | 493 | 160 | 3.0 | 0.08 |
| | Bladder Test | 0.5 | 0.2 | 762 | 160 | 4.7 | 0.03 |
| | Audio/Visual | -0.6 | 0.5 | 187 | 160 | 1.2 | 0.29 |
| 4 | Intercept | 11.7 | 1.2 | 11317 | 127 | 89.4 | <.001 |
| | Baseline Pelvic Pain | 0.1 | 0.1 | 65 | 127 | 0.5 | 0.48 |
| | QST | 0.3 | 0.1 | 801 | 127 | 6.3 | 0.01 |
| | Bladder Test | 0.4 | 0.3 | 262 | 127 | 2.1 | 0.15 |
| | Audio/Visual | 0.3 | 0.5 | 43 | 127 | 0.3 | 0.56 |

| | | | $\beta$ 95% CI | | | $\eta_p^2$ 95% CI | |
|---|---|---|---|---|---|---|---|
| Year | Parameter | $\beta$ | Low | High | $\eta_p^2$ | Low | High |
| 1 | Intercept | | -0.13 | 0.13 | 0.58 | 0.48 | 0.66 |
| | Baseline Pelvic Pain | 0.51 | 0.34 | 0.68 | 0.20 | 0.09 | 0.31 |
| | QST | 0.10 | -0.05 | 0.24 | 0.01 | 0.00 | 0.07 |
| | Bladder Test | 0.11 | -0.07 | 0.29 | 0.01 | 0.00 | 0.07 |
| | Audio/Visual | 0.03 | -0.11 | 0.17 | 0.00 | 0.00 | 0.04 |
| 2 | Intercept | | -0.16 | 0.16 | 0.52 | 0.41 | 0.62 |
| | Baseline Pelvic Pain | 0.26 | 0.05 | 0.47 | 0.05 | 0.00 | 0.14 |
| | QST | 0.10 | -0.08 | 0.27 | 0.01 | 0.00 | 0.07 |
| | Bladder Test | 0.19 | -0.03 | 0.41 | 0.02 | 0.00 | 0.10 |
| | Audio/Visual | 0.03 | -0.14 | 0.20 | 0.00 | 0.00 | 0.04 |
| 3 | Intercept | | -0.17 | 0.17 | 0.51 | 0.37 | 0.62 |
| | Baseline Pelvic Pain | 0.27 | 0.04 | 0.50 | 0.06 | 0.00 | 0.17 |
| | QST | 0.17 | -0.02 | 0.37 | 0.03 | 0.00 | 0.13 |
| | Bladder Test | 0.26 | 0.02 | 0.50 | 0.05 | 0.00 | 0.16 |
| | Audio/Visual | -0.10 | -0.29 | 0.09 | 0.01 | 0.00 | 0.09 |
| 4 | Intercept | | -0.19 | 0.19 | 0.52 | 0.37 | 0.63 |
| | Baseline Pelvic Pain | 0.10 | -0.18 | 0.38 | 0.01 | 0.00 | 0.08 |
| | QST | 0.27 | 0.06 | 0.49 | 0.07 | 0.00 | 0.20 |
| | Bladder Test | 0.21 | -0.08 | 0.51 | 0.02 | 0.00 | 0.12 |
| | Audio/Visual | 0.06 | -0.15 | 0.27 | 0.00 | 0.00 | 0.07 |

Table 3: **Regression Results for Summed Z-Score Sensory Constructs.** *Note.* The *df* for each model were: Year 1 (1, 138), Year 2 (1, 124), Year 3 (1, 92), Year 4 (1, 82).

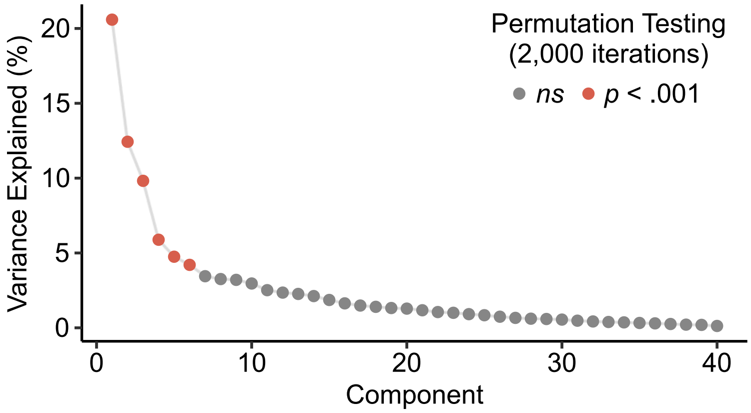


Figure 1: **Scree plot supports the first three principal components as important.** The first three components explain 43% of the variability in multimodal QST. Although components four, five, and six were significant via permutation testing, their geometric factor score plots were not interpretable and likely represented non-meaningful variation. Therefore, we chose to focus on the first three components that we interpreted as representing multimodal hypersensitivity (MMH), pressure pain threshold stimulus-response function (PPT-SR), and bladder pain hypersensitivity, respectively.

| PC | $\lambda_i$ | $\sigma^2$ (%) | Cumulated (%) | PC | $\lambda_i$ | $\sigma^2$ (%) | Cumulated (%) |
|---|---|---|---|---|---|---|---|
| 1* | 1639 | 20.6 | 21 | 21 | 93 | 1.2 | 90 |
| 2* | 990 | 12.4 | 33 | 22 | 84 | 1.1 | 91 |
| 3* | 782 | 9.8 | 43 | 23 | 80 | 1.0 | 92 |
| 4* | 468 | 5.9 | 49 | 24 | 73 | 0.9 | 93 |
| 5* | 378 | 4.8 | 53 | 25 | 67 | 0.8 | 94 |
| 6* | 335 | 4.2 | 58 | 26 | 59 | 0.7 | 95 |
| 7 | 275 | 3.4 | 61 | 27 | 53 | 0.7 | 95 |
| 8 | 259 | 3.3 | 64 | 28 | 49 | 0.6 | 96 |
| 9 | 255 | 3.2 | 68 | 29 | 47 | 0.6 | 96 |
| 10 | 236 | 3.0 | 71 | 30 | 44 | 0.5 | 97 |
| 11 | 200 | 2.5 | 73 | 31 | 38 | 0.5 | 97 |
| 12 | 187 | 2.4 | 75 | 32 | 34 | 0.4 | 98 |
| 13 | 180 | 2.3 | 78 | 33 | 31 | 0.4 | 98 |
| 14 | 169 | 2.1 | 80 | 34 | 29 | 0.4 | 99 |
| 15 | 148 | 1.9 | 82 | 35 | 26 | 0.3 | 99 |
| 16 | 130 | 1.6 | 83 | 36 | 24 | 0.3 | 99 |
| 17 | 118 | 1.5 | 85 | 37 | 20 | 0.3 | 99 |
| 18 | 112 | 1.4 | 86 | 38 | 17 | 0.2 | 100 |
| 19 | 105 | 1.3 | 88 | 39 | 16 | 0.2 | 100 |
| 20 | 102 | 1.3 | 89 | 40 | 10 | 0.1 | 100 |

Table 4: **Principal Component Eigenvalues ($\lambda_i$) and Explained Variances ($\sigma^2$) Obtained from CRAMPP QST Data Matrix.** *Note.* PC = principal component; * indicates $p$=.005 for 2,000 permutation iterations.

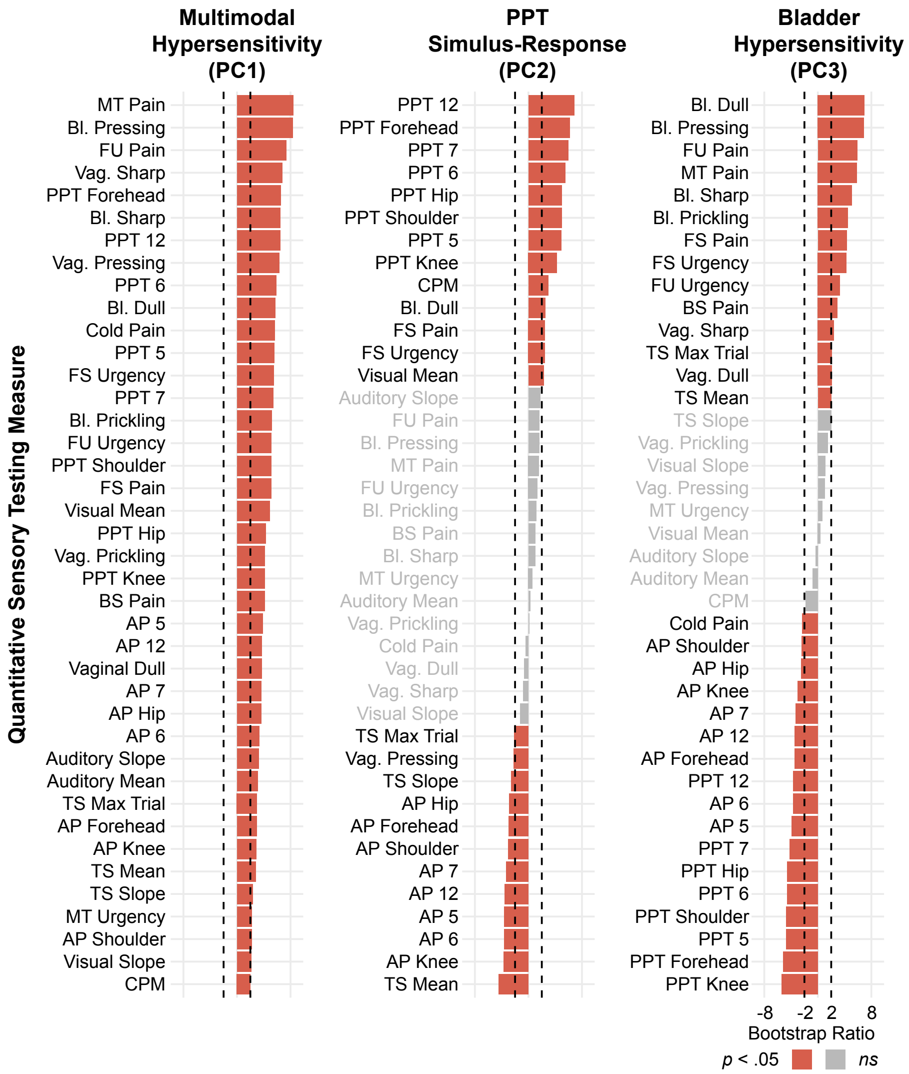


Figure 2: **Bootstrap ratios for each QST measure across the first three principal components (PCs).** A 2,000 iteration bootstrapping procedure calculated bootstrap ratios that are interpreted like Student's $t$-value and indicate the extent to which they significantly contributed to the variance of a PC. BS=baseline; FS=first sensation; FU=first urge; MT=max. tolerance; PPT=pressure pain threshold; TS=temporal summation; AP=after-pain; CPM=conditioned pain modulation; Bl.=bladder; Vag.=Vaginal; numbers denote clock face positions (e.g., 6=6 o'clock).

| Measure | *Mean* | *SD* | *Min* | *Max* |
|---|---|---|---|---|
| **PROMIS** (Total Score) | | | | |
| Pain Interference | 11.0 | 5.7 | 6 | 29 |
| Pain Behavior | 18.3 | 7.7 | 7 | 35 |
| Depression | 15.6 | 6.7 | 8 | 37 |
| Anxiety | 17.2 | 6.0 | 7 | 35 |
| **Menstrual Pain (0-100 VAS)** | | | | |
| Without NSAIDs | 64.8 | 25.8 | 0 | 100 |
| **Interstitial Cystitis** | | | | |
| Symptom Index (ICSI) | 5.7 | 4.1 | 0 | 18 |
| Problem Index (ICPI) | 3.5 | 3.8 | 0 | 15 |
| **GUPI** | | | | |
| Urinary | 3.5 | 2.7 | 0 | 10 |
| Quality of Life | 2.9 | 2.9 | 0 | 12 |
| Pain | 5.2 | 5.2 | 0 | 20 |
| Total | 11.6 | 9.6 | 0 | 41 |
| **CMSI (for 3 months)** | | | | |
| During the last year | 5.4 | 6.7 | 0 | 30 |
| During your lifetime | 4.8 | 6.6 | 0 | 34 |
| **Global Mental Health** | 3.7 | 0.9 | 1 | 5 |
| **Global Physical Health** | 3.4 | 0.9 | 1 | 5 |
| **BSI (Somatic Symptoms)** | 3.1 | 3.4 | 0 | 21 |

Table 5: **Descriptive Statistics of Self-Report Measures.** *Note.* $n$=200 for all measures except menstrual pain ($n$=194) and global mental/physical health ($n$=199); Global mental/physical health have opposite directionality (i.e., increased score denotes better health); PROMIS=Patient Reported Outcomes Measurement Information System; NSAIDs=Non-Steroidal Anti-Inflammatory Drugs; GUPI=Genitourinary Pain Index; CMSI=Complex Medical Symptoms Inventory; BSI=Brief Symptom Inventory.

| | Year | | | | |
|---|---|---|---|---|---|
| | Baseline | 1 | 2 | 3 | 4 |
| *Mean* | 15.6 | 15.5 | 13.5 | 12.4 | 11.0 |
| *SD* | 18.3 | 17.8 | 15.1 | 14.9 | 12.6 |
| *Min - Max* | 0 - 77 | 0 - 95 | 0 - 68 | 0 - 72 | 0 - 54 |
| *Median* | 7.8 | 9.3 | 8.0 | 7.0 | 5.7 |
| $n$ | 200 | 143 | 129 | 97 | 87 |

Table 6: **Descriptive Statistics of Pelvic Pain Outcome.** *Note.* CI=Confidence Interval; LL=Lower Level; UL=Upper Level

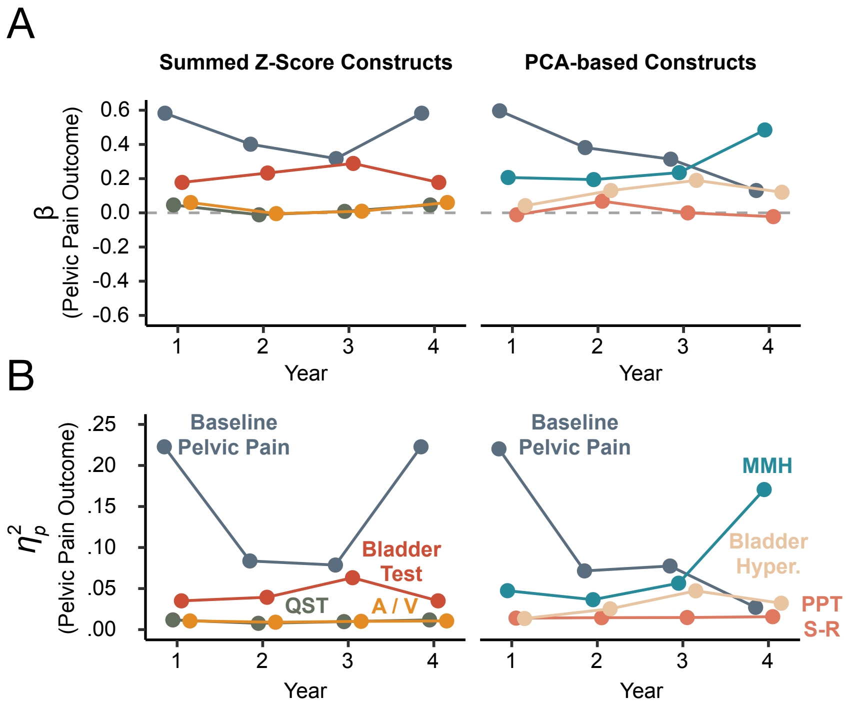


Figure 3: **Adjusting for population prevalence rates replicated the observed sample-wise regression results across both construct models.** A) Standardized regression coefficients ($\beta$'s) are plotted across the collected annual questionnaire period for both the summed Z-score constructs (left) and the PCA-based constructs (right). All $\beta$ values are approximately zero or positive, indicating in general that increased sensitivity resulted in worse pelvic pain outcome. See B for colored labels denoting predictor variables. B) Unique explained variance in pelvic pain outcome as a function of each predictor ($\eta_p^2$) is plotted across the collected annual questionnaire period for both the summed Z-score constructs (left) and the PCA-based constructs (right). When comparing to the unadjusted regression results, the pattern of change over time is similar, especially in the case of the PCA-based constructs. Namely, MMH increases in predictive strength of pelvic pain outcome over time, while the opposite trend is observed with baseline pelvic pain.; A/V = Audio/Visual.

| Year | Parameter | $b$ | $SE$ | $SS$ | $MSE$ | $F$ | $p$ |
|---|---|---|---|---|---|---|---|
| 1 | Intercept | 16.3 | 1.2 | 37790.9 | 193.7 | 195.1 | <.001 |
| | Baseline Pelvic Pain | 0.6 | 0.1 | 7286.7 | 193.7 | 37.6 | <.001 |
| | PC 1 - MMH | 1.1 | 0.5 | 877.8 | 193.7 | 4.5 | 0.04 |
| | PC 2 - PPT S-R | -0.3 | 0.6 | 46.3 | 193.7 | 0.2 | 0.63 |
| | PC 3 - Bladder Hyper. | -0.4 | 0.7 | 61.3 | 193.7 | 0.3 | 0.57 |
| 2 | Intercept | 14.3 | 1.2 | 25600.3 | 186.3 | 137.4 | <.001 |
| | Baseline Pelvic Pain | 0.3 | 0.1 | 1191.6 | 186.3 | 6.4 | 0.01 |
| | PC 1 - MMH | 1.4 | 0.5 | 1314.5 | 186.3 | 7.1 | 0.01 |
| | PC 2 - PPT S-R | 0.1 | 0.6 | 1.3 | 186.3 | 0.0 | 0.93 |
| | PC 3 - Bladder Hyper. | 0.2 | 0.7 | 9.8 | 186.3 | 0.1 | 0.82 |
| 3 | Intercept | 12.6 | 1.3 | 14737.4 | 160.3 | 91.9 | <.001 |
| | Baseline Pelvic Pain | 0.3 | 0.1 | 1127.8 | 160.3 | 7.0 | 0.01 |
| | PC 1 - MMH | 1.6 | 0.6 | 1365.8 | 160.3 | 8.5 | 0.004 |
| | PC 2 - PPT S-R | -0.7 | 0.6 | 281.8 | 160.3 | 1.8 | 0.19 |
| | PC 3 - Bladder Hyper. | 0.8 | 0.7 | 213.0 | 160.3 | 1.3 | 0.25 |
| 4 | Intercept | 11.8 | 1.2 | 11505.9 | 126.7 | 90.8 | <.001 |
| | Baseline Pelvic Pain | 0.1 | 0.1 | 49.1 | 126.7 | 0.4 | 0.54 |
| | PC 1 - MMH | 2.0 | 0.5 | 1762.8 | 126.7 | 13.9 | <.001 |
| | PC 2 - PPT S-R | -0.3 | 0.6 | 31.2 | 126.7 | 0.2 | 0.62 |
| | PC 3 - Bladder Hyper. | 0.2 | 0.7 | 10.9 | 126.7 | 0.1 | 0.77 |

| | | | $\beta$ 95% CI | | | $\eta_p^2$ 95% CI | |
|---|---|---|---|---|---|---|---|
| Year | Parameter | $\beta$ | Low | High | $\eta_p^2$ | Low | High |
| 1 | Intercept | | | | 0.59 | 0.48 | 0.66 |
| | Baseline Pelvic Pain | 0.54 | 0.36 | 0.71 | 0.21 | 0.11 | 0.33 |
| | PC 1 - MMH | 0.17 | 0.01 | 0.33 | 0.03 | 0.00 | 0.11 |
| | PC 2 - PPT S-R | -0.03 | -0.16 | 0.10 | <.01 | 0.00 | 0.04 |
| | PC 3 - Bladder Hyper. | -0.04 | -0.19 | 0.10 | <.01 | 0.00 | 0.04 |
| 2 | Intercept | | | | 0.53 | 0.41 | 0.62 |
| | Baseline Pelvic Pain | 0.27 | 0.06 | 0.48 | 0.05 | 0.00 | 0.14 |
| | PC 1 - MMH | 0.25 | 0.06 | 0.44 | 0.05 | 0.00 | 0.15 |
| | PC 2 - PPT S-R | 0.01 | -0.16 | 0.18 | <.01 | 0.00 | 0.02 |
| | PC 3 - Bladder Hyper. | 0.02 | -0.16 | 0.20 | <.01 | 0.00 | 0.03 |
| 3 | Intercept | | | | 0.50 | 0.36 | 0.61 |
| | Baseline Pelvic Pain | 0.31 | 0.08 | 0.54 | 0.07 | 0.00 | 0.19 |
| | PC 1 - MMH | 0.31 | 0.10 | 0.51 | 0.08 | 0.01 | 0.21 |
| | PC 2 - PPT S-R | -0.12 | -0.30 | 0.06 | 0.02 | 0.00 | 0.11 |
| | PC 3 - Bladder Hyper. | 0.11 | -0.08 | 0.30 | 0.01 | 0.00 | 0.10 |
| 4 | Intercept | | | | 0.53 | 0.38 | 0.64 |
| | Baseline Pelvic Pain | 0.09 | -0.19 | 0.36 | <.01 | 0.00 | 0.07 |
| | PC 1 - MMH | 0.44 | 0.21 | 0.68 | 0.15 | 0.03 | 0.29 |
| | PC 2 - PPT S-R | -0.05 | -0.27 | 0.16 | <.01 | 0.00 | 0.07 |
| | PC 3 - Bladder Hyper. | 0.03 | -0.20 | 0.26 | <.01 | 0.00 | 0.05 |

Table 7: **Regression Results for PCA-based Construct Models.** *Note.* The *df* for each model were: Year 1 (1, 138), Year 2 (1, 124), Year 3 (1, 92), Year 4 (1, 82).

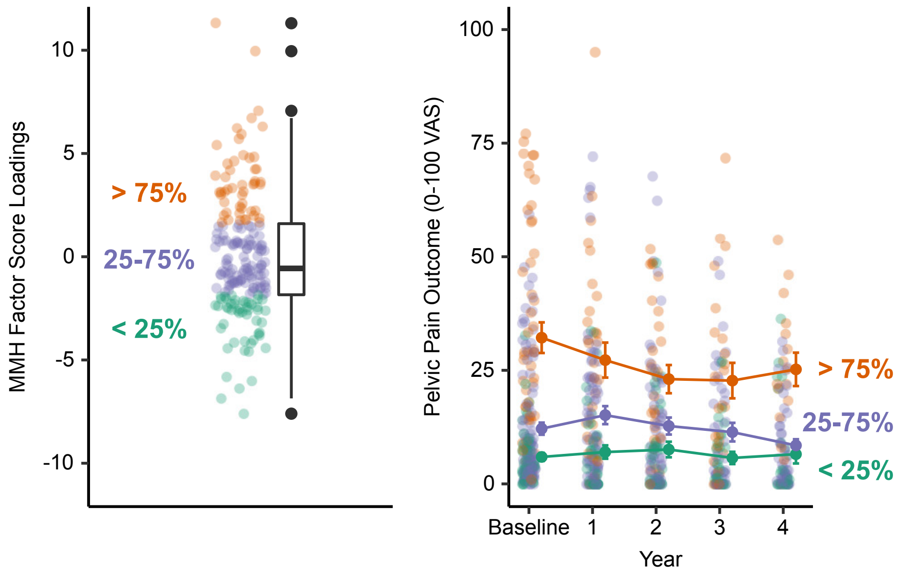


Figure 4: **MMH Outer-Quartile Participants Demonstrate Markedly Different Pelvic Pain Trajectories.** Participants were first differentiated based on whether they are inside or outside the inter-quartile range for baseline MMH factor loadings (left). Participants with MMH factor loadings > 75% of the sample demonstrated worse pelvic pain outcome at baseline that persisted across the four-year follow-up period (right). Error bars are standard error of the mean.

#### References


Aslam, N., Harrison, G., Khan, K., & Patwardhan, S. (2009). Visceral hyperalgesia in chronic pelvic pain. *BJOG: An International Journal of Obstetrics & Gynaecology*, *116*(12), 1551–1555. https://doi.org/10.1111/j.1471-0528.2009.02305.x

Beaton, D., Chin Fatt, C. R., & Abdi, H. (2014). An ExPosition of multivariate analysis with the singular value decomposition in r. *Computational Statistics & Data Analysis*, *72*, 176–189. https://doi.org/10.1016/j.csda.2013.11.006

Beissner, F., Brandau, A., Henke, C., Felden, L., Baumgärtner, U., Treede, R.-D., Oertel, B. G., & Lötsch, J. (2010). Quick discrimination of adelta and c fiber mediated pain based on three verbal descriptors (K. Ikeda, Ed.). *PLoS ONE*, *5*(9), e12944. https://doi.org/10.1371/journal.pone.0012944

Ben-Shachar, M. S., Makowski, D., & Lüdecke, D. (2020). *Compute and interpret indices of effect size*. https://github.com/easystats/effectsize

Broderick, J., DeWit, E. M., Rothrock, N., Crane, P., & Forrest, C. B. (2013). Advances in patient reported outcomes: The NIH PROMIS measures [Number: 1 Publisher: Ubiquity Press]. *eGEMs (Generating Evidence & Methods to improve patient outcomes)*, *1*(1), 12. https://doi.org/10.13063/2327-9214.1015

Cathcart, S., Winefield, A. H., Rolan, P., & Lushington, K. (2009). Reliability of temporal summation and diffuse noxious inhibitory control [Publisher: Hindawi]. *Pain Research and Management*, *14*(6), 433–438. https://doi.org/10.1155/2009/523098

Clemens, J. Q., Calhoun, E. A., Litwin, M. S., McNaughton-Collins, M., Kusek, J. W., Crowley, E. M., & Landis, J. R. (2009). Validation of a modified national institutes of health chronic prostatitis symptom index to assess genitourinary pain in both men and women. *Urology*, *74*(5), 983–987.e3. https://doi.org/10.1016/j.urology.2009.06.078

Derogatis, L. R., & Melisaratos, N. (1983). The brief symptom inventory: An introductory report. *Psychological Medicine*, *13*(3), 595–605. https://doi.org/10.1017/S0033291700048017

D'haenens, W., Dhooge, I., Maes, L., Bockstael, A., Keppler, H., Philips, B., Swinnen, F., & Vinck, B. M. (2009). The clinical value of the multiple-frequency 80-hz auditory steady-state response in adults with normal hearing and hearing loss. *Archives of Otolaryngology–Head & Neck Surgery*, *135*(5), 496–506. https://doi.org/10.1001/archoto.2009.32

FitzGerald, M. P., Koch, D., & Senka, J. (2005). Visceral and cutaneous sensory testing in patients with painful bladder syndrome. *Neurourology and Urodynamics*, *24*(7), 627–632. https://doi.org/10.1002/nau.20178

Friedman, D. I., & De Ver Dye, T. (2009). Migraine and the environment. *Headache: The Journal of Head and Face Pain*, *49*(6), 941–952. https://doi.org/10.1111/j.1526-4610.2009.01443.x

Gracely, R. H., & Kwilosz, D. M. (1988). The descriptor differential scale: Applying psychophysical principles to clinical pain assessment: *Pain*, *35*(3), 279–288. https://doi.org/10.1016/0304-3959(88)90138-8

Granges, G., & Littlejohn, G. (1993). Pressure pain threshold in pain-free subjects, in patients with chronic regional pain syndromes, and in patients with fibromyalgia syndrome. *Arthritis & Rheumatism*, *36*(5), 642–646. https://doi.org/10.1002/art.1780360510

Hanno, P. M., Burks, D. A., Clemens, J. Q., Dmochowski, R. R., Erickson, D., FitzGerald, M. P., Forrest, J. B., Gordon, B., Gray, M., Mayer, R. D., Newman, D., Nyberg, L., Payne, C. K., Wesselmann, U., & Faraday, M. M. (2011). AUA guideline for the diagnosis and treatment of interstitial cystitis/bladder pain syndrome. *Journal of Urology*, *185*(6), 2162–2170. https://doi.org/10.1016/j.juro.2011.03.064

Harte, S. E., Ichesco, E., Hampson, J. P., Peltier, S. J., Schmidt-Wilcke, T., Clauw, D. J., & Harris, R. E. (2016). Pharmacologic attenuation of cross-modal sensory augmentation within the chronic pain insula: *PAIN*, *157*(9), 1933–1945. https://doi.org/10.1097/j.pain.0000000000000593

Haylen, B. T., de Ridder, D., Freeman, R. M., Swift, S. E., Berghmans, B., Lee, J., Monga, A., Petri, E., Rizk, D. E., Sand, P. K., & Schaer, G. N. (2010). An international urogynecological association (IUGA)/international continence society (ICS) joint report on the terminology for female pelvic floor dysfunction: Terminology for female pelvic floor dysfunction. *Neurourology and Urodynamics*, *29*(1), 4–20. https://doi.org/10.1002/nau.20798

Hellman, K. M., Datta, A., Steiner, N. D., Kane Morlock, J. N., Garrison, E. F., Clauw, D. J., & Tu, F. F. (2018). Identification of experimental bladder sensitivity among dysmenorrhea sufferers. *American Journal of Obstetrics and Gynecology*, *219*(1), 84.e1–84.e8. https://doi.org/10.1016/j.ajog.2018.04.030

Hellman, K. M., Patanwala, I. Y., Pozolo, K. E., & Tu, F. F. (2015). Multimodal nociceptive mechanisms underlying chronic pelvic pain. *American Journal of Obstetrics and Gynecology*, *213*(6), 827.e1–827.e9. https://doi.org/10.1016/j.ajog.2015.08.038

Hellman, K. M., Roth, G. E., Dillane, K. E., Garrison, E. F., Oladosu, F. A., Clauw, D. J., & Tu, F. F. (2020). Dysmenorrhea subtypes exhibit differential quantitative sensory assessment profiles. *PAIN*, *161*(6), 1227–1236. https://doi.org/10.1097/j.pain.0000000000001826

Henderson, E. (2020). *Ghibli: Studio ghibli colour palettes* (Version 0.3.2). https://CRAN.R-project.org/package=ghibli

Hollins, M., Harper, D., Gallagher, S., Owings, E. W., Lim, P. F., Miller, V., Siddiqi, M. Q., & Maixner, W. (2009). Perceived intensity and unpleasantness of cutaneous and auditory stimuli: An evaluation of the generalized hypervigilance hypothesis: *Pain*, *141*(3), 215–221. https://doi.org/10.1016/j.pain.2008.10.003

Iacovides, S., Avidon, I., & Baker, F. C. (2015). What we know about primary dysmenorrhea today: A critical review. *Human Reproduction Update*, *21*(6), 762–778. https://doi.org/10.1093/humupd/dmv039

Kanazawa, M., Hongo, M., & Fukudo, S. (2011). Visceral hypersensitivity in irritable bowel syndrome. *Journal of Gastroenterology and Hepatology*, *26*, 119–121. https://doi.org/10.1111/j.1440-1746.2011.06640.x

Kmiecik, M. J., Tu, F. F., Silton, R. L., Dillane, K. E., Roth, G. E., Harte, S. E., & Hellman, K. M. (2021). Cortical mechanisms of visual hypersensitivity in women at risk for chronic pelvic pain. *PAIN*. https://doi.org/10.1097/j.pain.0000000000002469

López-Solà, M., Pujol, J., Wager, T. D., Garcia-Fontanals, A., Blanco-Hinojo, L., Garcia-Blanco, S., Poca-Dias, V., Harrison, B. J., Contreras-Rodríguez, O., Monfort, J., Garcia-Fructuoso, F., & Deus, J. (2014). Altered functional magnetic resonance imaging responses to nonpainful sensory stimulation in fibromyalgia patients: Brain response to nonpainful multisensory stimulation in fibromyalgia. *Arthritis & Rheumatology*, *66*(11), 3200–3209. https://doi.org/10.1002/art.38781

Martenson, M. E., Halawa, O. I., Tonsfeldt, K. J., Maxwell, C. A., Hammack, N., Mist, S. D., Pennesi, M. E., Bennett, R. M., Mauer, K. M., Jones,

K. D., & Heinricher, M. M. (2016). A possible neural mechanism for photosensitivity in chronic pain. *PAIN*, *157*(4), 868–878. https://doi.org/10.1097/j.pain.0000000000000450

Neuwirth, E. (2014). *RColorBrewer: ColorBrewer palettes*. R package version 1.1-2. https://CRAN.R-project.org/package=RColorBrewer

Nir, R.-R., & Yarnitsky, D. (2015). Conditioned pain modulation. *Current Opinion in Supportive and Palliative Care*, *9*(2), 131–137. https://doi.org/10.1097/SPC.0000000000000126

O'Brien, A. T., Deitos, A., Triñanes Pego, Y., Fregni, F., & Carrillo-de-la-Peña, M. T. (2018). Defective endogenous pain modulation in fibromyalgia: A meta-analysis of temporal summation and conditioned pain modulation paradigms. *The Journal of Pain*, *19*(8), 819–836. https://doi.org/10.1016/j.jpain.2018.01.010

O'Leary, M. P., Sant, G. R., Fowler, F. J., Whitmore, K. E., & Spolarich-Kroll, J. (1997). The interstitial cystitis symptom index and problem index. *Urology*, *49*(5), 58–63. https://doi.org/https://doi.org/10.1016/S0090-4295(99)80333-1

Payne, L. A., Rapkin, A., Seidman, L., Zeltzer, L., & Tsao, J. (2017). Experimental and procedural pain responses in primary dysmenorrhea: A systematic review. *Journal of Pain Research*, *Volume 10*, 2233–2246. https://doi.org/10.2147/JPR.S143512

Payne, L. A., Seidman, L. C., Sim, M.-S., Rapkin, A. J., Naliboff, B. D., & Zeltzer, L. K. (2019). Experimental evaluation of central pain processes in young women with primary dysmenorrhea: *PAIN*, *160*(6), 1421–1430. https://doi.org/10.1097/j.pain.0000000000001516

Picou, E. M., Ricketts, T. A., & Hornsby, B. W. Y. (2013). How hearing aids, background noise, and visual cues influence objective listening effort: *Ear and Hearing*, *34*(5), e52–e64. https://doi.org/10.1097/AUD.0b013e31827f0431

Revelle, W. (2020). *Psych: Procedures for personality and psychological research* (Version 2.0.12). Northwestern University, Evanston, IL. https://CRAN.R-project.org/package=psych

Thompson, H. D., Tang, S., & Jarrell, J. F. (2020). Temporal summation in chronic pelvic pain. *Journal of Obstetrics and Gynaecology Canada*, *42*(5), 556–560. https://doi.org/10.1016/j.jogc.2019.09.012

Tu, F. F., Epstein, A. E., Pozolo, K. E., Sexton, D. L., Melnyk, A. I., & Hellman, K. M. (2013). A noninvasive bladder sensory test supports a role for dysmenorrhea increasing bladder noxious mechanosensitivity. *The Clinical Journal of Pain*, *29*(10), 883–890. https://doi.org/10.1097/AJP.0b013e31827a71a3

Tu, F. F., Kane, J. N., & Hellman, K. M. (2017). Noninvasive experimental bladder pain assessment in painful bladder syndrome. *BJOG: An International Journal of Obstetrics & Gynaecology*, *124*(2), 283–291. https://doi.org/10.1111/1471-0528.14433

Westling, A. M., Tu, F. F., Griffith, J. W., & Hellman, K. M. (2013). The association of dysmenorrhea with noncyclic pelvic pain accounting for psychological factors. *American Journal of Obstetrics and Gynecology*, *209*(5), 422.e1–422.e10. https://doi.org/10.1016/j.ajog.2013.08.020

Wickham, H., Averick, M., Bryan, J., Chang, W., McGowan, L. D., François, R., Grolemund, G., Hayes, A., Henry, L., Hester, J., Kuhn, M., Pedersen, T. L., Miller, E., Bache, S. M., Müller, K., Ooms, J., Robinson, D., Seidel, D. P., Spinu, V., . . . Yutani, H. (2019). Welcome to the tidyverse. *Journal of Open Source Software*, *4*(43), 1686. https://doi.org/10.21105/joss.01686

Wilbarger, J. L., & Cook, D. B. (2011). Multisensory hypersensitivity in women with fibromyalgia: Implications for well being and intervention. *Archives of Physical Medicine and Rehabilitation*, *92*(4), 653–656. https://doi.org/10.1016/j.apmr.2010.10.029

Williams, D. A., & Schilling, S. (2009). Advances in the assessment of fibromyalgia. *Rheumatic Disease Clinics of North America*, *35*(2), 339–357. https://doi.org/10.1016/j.rdc.2009.05.007

Wolfe, F., Smythe, H. A., Yunus, M. B., Bennett, R. M., Bombardier, C., Goldenberg, D. L., Tugwell, P., Campbell, S. M., Abeles, M., Clark, P., Fam, A. G., Farber, S. J., Fiechtner, J. J., Michael Franklin, C., Gatter, R. A., Hamaty, D., Lessard, J., Lichtbroun, A. S., Masi, A. T., . . . Sheon, R. P. (1990). The american college of rheumatology 1990 criteria for the classification of fibromyalgia. *Arthritis & Rheumatism*, *33*(2), 160–172. https://doi.org/10.1002/art.1780330203

Yarnitsky, D., Crispel, Y., Eisenberg, E., Granovsky, Y., Ben-Nun, A., Sprecher, E., Best, L.-A., & Granot, M. (2008). Prediction of chronic post-operative pain: Pre-operative DNIC testing identifies patients at risk. *Pain*, *138*(1), 22–28. https://doi.org/10.1016/j.pain.2007.10.033

Yarnitsky, D., Granot, M., Nahman-Averbuch, H., Khamaisi, M., & Granovsky, Y. (2012). Conditioned pain modulation predicts duloxetine efficacy in painful diabetic neuropathy. *Pain*, *153*(6), 1193–1198. https://doi.org/10.1016/j.pain.2012.02.021
